# Aneuploidy and structural genome complexity beyond the EWSR1 fusion is associated with evolution and outcome in clear cell sarcoma

**DOI:** 10.64898/2026.11.20.26368414

**Authors:** Akanksha Farswan, Christopher Davies, Dawid Krzeciesa, Boyu Yu, Wenxin Chao, Runtian Lin, Hongtao Ye, Thomas Butters, Emine Hapitoglu, Benjamin M. Chain, Simon Haefliger, Laura Da Silva Tavares, Sidi Yang, Abigail Bunkum, Rija Zaidi, Shadi Hames-Fathi, Hazem Moussa, Samira Nur, Daniel Hughes, Sumathi Vajipuri, Simone Zaccaria, Mahbubl Ahmed, Fernanda Amary, Roberto Tirabosco, Christopher D. Steele, Sara Waise, Tom Lesluyes, Christian Brandts, Hanno Glimm, Stefan Fröhling, Thomas Kindler, Carolin Mogler, Annika Baude-Müller, Daniel Baumhoer, Peter Van Loo, Adrienne M. Flanagan, Sandra J. Strauss, Priya Chudasama, Sebastian Bauer, Nischalan Pillay, the Clear Cell Sarcoma Consortium

**Author notes:** **Corresponding authors:** Prof. Nischalan Pillay; Research Department of Pathology, UCL Cancer Institute, London, United, Kingdom; Royal National Orthopedic Hospital Trust, London, United Kingdom, Prof. Sebastian Bauer; Department of Medical Oncology and Sarcoma Center, West German Cancer Center, University Duisburg-Essen, Medical School, Essen, Germany; DKTK partner site Essen, German Cancer Consortium (DKTK), Heidelberg, Germany. These authors contributed equally as first authors (Akanksha Farswan, Christopher Davies, Dawid Krzeciesa). These authors jointly supervised this work (Sebastian Bauer, Nischalan Pillay).

## Abstract

Clear cell sarcoma (CCS) is an ultra-rare cancer defined by EWSR1-CREB gene family fusions with clinical outcomes that range from indolent to chemotherapy-refractory metastatic disease. Through multi-omic profiling of primary, recurrent and metastatic tumours and cell lines, we show that CCS is more genomically complex than previously recognised. Beyond the fusion, tumours acquire chromoplexy, chromothripsis and an intermediate aneuploidy burden, with ongoing subclonal diversification. Recurrent events include chromosome17q gain and inactivation of the *MYC* antagonist *MGA*. Enriched transcriptional signatures of chromosomal instability,17q gain, 9p21.3 loss and *TERT* alteration each double the hazard of death, supporting a new molecular based CCS risk stratification framework at diagnosis. Transcriptomes converge on a MYC-driven ribosome biogenesis programme, a previously unrecognised therapeutic vulnerability. These findings address a central paradox of CCS, a shared oncogenic identity with divergent clinical fate, implicating tolerated aneuploidy and chromosomal diversification as candidate determinants of progression.

## Introduction

Clear cell sarcoma of soft tissue (CCS) is an ultra-rare, aggressive malignancy of deep soft tissues predominantly affecting young adults (Enzinger, 1965) (Graadt Van Roggen et al., 1998) (Chung & Enzinger, 1983) most commonly presenting in the lower extremities and shares morphologic and immunohistochemical features with melanoma. A pathognomonic fusion, *EWSR1::ATF1* t(12;22)(q13;q12), is the defining hallmark of clear cell sarcoma. This involves a translocation of the 5’ region of the *EWSR1* gene with the 3’ region of the activating transcription factor 1 (*ATF1*) gene on chromosome 12. *ATF1* encodes an activating transcription factor that belongs to the cAMP response element-binding protein (CREB)/ATF family of basic leucine zipper (bZIP) transcription factors (Chowdhury et al., 2024). The *EWSR1::ATF1* fusion hence joins the transcription activation domain of *EWSR1* with the DNA-binding domain of *ATF1* or rarely other members of the cAMP family (*CREB1* or *CREM*). The fusion occupies the microphthalmia-associated transcription factor (MITF) promoter, drives MITF expression which mediates CCS cell survival and proliferation and confers the melanocytic phenotype that underlies the histological resemblance to melanoma and complicates the morphological differential diagnosis (Davis et al., 2006) (Antonescu et al., 2002). EWSR1::ATF1 binds predominantly distal regulatory elements and co-opts the melanocytic core regulatory circuit, MITF, SOX10 and TFAP2A onto fusion specific enhancers, such that fusion depletion releases these factors, induces melanosomal genes (MLANA, TYRP1, PMEL) and increases melanosome number (Möller et al., 2022).

The fusion therefore specifies melanocytic identity while restricting terminal differentiation. Moreover, *EWSR1::ATF1* is sufficient to induces CCS in mouse models (Panza et al., 2021) and the differentiation programmes that emerge on its removal implicate neural crest-derived precursors as putative cells of origin (Antonescu et al., 2002) (Davis et al., 2006) (Panza et al., 2021) (Möller et al., 2022). CCS, like other fusion-driven sarcomas, is often regarded as genomically “simple” with a balanced chromosomal and an overall diploid profile with low tumour mutational burden (J. Li et al., 2021) (Panza et al., 2021) (Rasmussen et al., 2023). Recurrent copy number alterations (CNAs) include arm-level or whole chromosome gains of chromosomes 7, 8 and 1q, and deletions on chromosome 9 (*CDKN2A*) and 19 (Panza et al., 2021) (Rasmussen et al., 2023) (J. Li et al., 2021).

CCS carries a high risk of local recurrence and synchronous metastasis with rates reported as high as 84% for local recurrence (Ibrahim et al., 2018). 5-year overall survival rates are reported to be around 51% with limited treatment options (A. B. Li et al., 2019) (H. Y. Wang et al., 2025). Deciphering a comprehensive somatic genomic landscape of CCS that could identify putative therapeutic targets remains a challenge due to its ultra-rarity and lack of available high-quality frozen material with paired germline samples. Previous genomic studies have been conducted on cohorts using unpaired low-pass whole genome or exome data from unpaired formalin-fixed paraffin embedded (FFPE) tissue (Lee et al., 2021) (Rasmussen et al., 2023), which are prone to germline artefacts in mutation calling and offer limited power and resolution to precisely infer genomic structural variation. Consequently, critical questions concerning the spectrum of structural variants in CCS and any potential genomic subgroup that could assist in identifying therapeutic opportunities are yet to be adequately resolved. CCS shows low response rates to conventional soft tissue sarcoma chemotherapy regimens (Jones et al., 2011); and targeted approaches such as MET inhibition have shown only modest activity. Management of localised disease therefore relies on wide surgical resection with negative margins, with radiotherapy for local control in selected cases and consideration of regional nodal assessment given the propensity for lymphatic spread. Despite being defined by a common oncogenic driver, CCS exhibits striking clinical heterogeneity, ranging from indolent localised disease to rapidly progressive, chemotherapy-refractory metastatic dissemination (Kawai et al., 2007). The molecular basis of this divergence remains largely unknown.

Resolving this paradox of CCS sharing a near universal genomic and transcriptional identity yet divergent clinical behaviour requires moving beyond the initiating fusion to understand the evolutionary processes that generate biological diversity within the disease. Here, we performed comprehensive integrative profiling of a large multinational cohort to broaden the single-driver view of CCS biology and uncover the genomic and transcriptional programs that underpin tumour progression, metastatic competence, and therapeutic resistance. In addition to identifying the basis of genomic heterogeneity, molecular correlates of adverse outcome, and potential “druggable” pathways, the data from this study serve as resource for translational research, enabling future mechanistic studies to be designed, informing biologically-driven patient stratification, and providing a framework for the development of biomarker-guided clinical trials in this rare and understudied cancer.

## Results

### Clinical characteristics and mutational landscape of CCS

We analysed a multi-institutional cohort of 136 CCS individuals of which 61 had fresh-frozen tumour samples with matched normal DNA available, constituting the discovery cohort (Figure 1A and B). The Germany cohort also included gastro-intestinal CCS cases (GI-CCS) (n = 8). The samples covered all clinical stages of the disease (primary, locally recurrent and metastatic tumours) (Supplementary Table 1) with multi-regional and longitudinal sampling of primary and metastatic lesions from a patient with a representative treatment history and clinical course. Ten CCS cell lines (Figure 1A) were additionally profiled to establish their fidelity as preclinical models of CCS. Three cell lines (MUG-Lucifer-prim, MUG-Lucifer-met (Karner et al., 2023) and CCS-SN1) had a paired matching normal sample for somatic mutation calling. Samples underwent deep molecular characterization including whole genome or exome, mRNA sequencing, methylation array and T-cell receptor repertoire profiling alongside curated clinical metadata (Figure 1C). Unpaired samples including snap frozen tumour, FFPE or external sequencing data from an additional 33 individuals constituted the expansion cohort where more focussed molecular testing was carried out.

**Figure 1:**
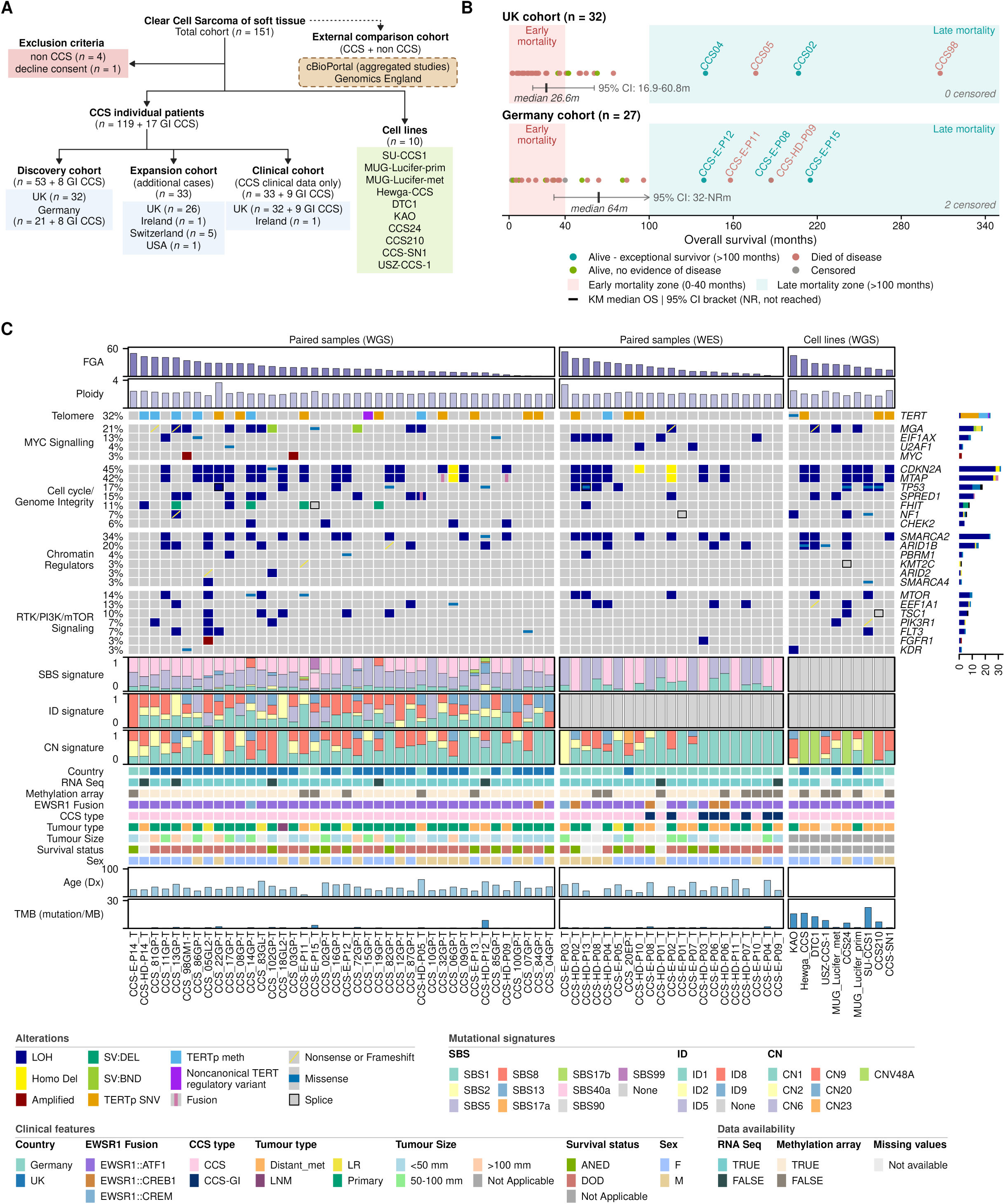
Clinical characteristics and mutational landscape of CCS A: CONSORT-style diagram showing patient inclusion, cohort allocation, and external datasets used for genomic, transcriptomic, and clinical analyses. Sample numbers are indicated throughout. B: Distribution of overall survival in the UK (upper strip) and Germany (lower strip) cohorts: Each dot represents one patient, plotted by overall survival (OS) in months and coloured by outcome (teal, alive with OS >100 months; green, alive with no evidence of disease; muted red, died of disease; grey, censored or lost to follow-up). Shaded zones denote early mortality (0–40 months) and exceptional survivors (>100 months). Case IDs are shown for patients surviving >100 months. Beneath each strip, the vertical black bar shows the Kaplan–Meier estimate of median OS and the horizontal bracket shows its 95% confidence interval (CI). Death from disease was the event; patients alive at last follow-up or lost to follow-up were censored at their last contact. Median OS was 26.6 months (95% CI 16.9–60.8) in the UK cohort and 64 months (95% CI 32–NR) in the Germany cohort. An arrow indicates an upper confidence limit that was not reached (NR). The number of censored patients in each cohort is shown at the top right of each strip. C: Oncoprint summarising genomic alterations, mutational signatures and clinical features across the clear cell sarcoma (CCS) cohort, including primary tumours, local recurrences, metastatic lesions, and established cell lines, with each column representing an individual tumour. The top panels show the fraction of the genome altered (FGA, %) and tumour ploidy for each sample. These are followed by the telomere maintenance status-orange/purple for TERT promoter mutation and blue for TERT hypermethylation and by genomic alterations in key cancer genes, grouped by pathways: MYC signalling, cell cycle/genome integrity, chromatin regulators, and RTK/PI3K/mTOR signalling. Alterations include copy-number changes, structural variants, gene fusions, promoter alterations, and coding mutations, as indicated in the key and the percentage of samples altered is shown on the left. The middle panels show the relative contribution of single-base substitution (SBS), small insertion/deletion (ID) and copy-number (CN) mutational signatures in each sample. The bottom panels show sample and clinical annotations: country of origin, availability of RNA sequencing and methylation array data, EWSR1 fusion partner (ATF1/CREB1/CREM), CCS type, tumour type (primary/recurrence/metastasis), tumour size, survival status, sex, age at diagnosis (years), and tumour mutational burden (TMB, mutations/Mb). Samples are ordered by genomic complexity and grouped according to paired specimens and cell lines.

Furthermore, samples with only clinical data constituted the CCS clinical data only cohort (Figure 1A). All patient samples from the discovery and expansion cohort harboured the driver fusion identified through either standard of care clinical testing or through the current research study. The median age at primary diagnosis was 32 years (range: 5 to 84 years) and median tumour size at primary diagnosis was 40 mm (range: 6 to 300mm) with an outlier case with 700mm primary tumour size involving an entire leg (Supplementary Table 1). The most common primary sites were the extremities (leg, n = 40; foot/ankle, n = 33; hand, n=24); 33.58% (n = 44) of individuals presented with either lymphatic (n=16) or systemic (n = 28) synchronous metastasis at the time of primary diagnosis (Supplementary Table 1). Follow-up data were available for 102 of 136 individuals, with a median follow-up of 32 months; of these, 35 (34.3%) were alive with or without evidence of disease at last follow-up. Primary tumours presenting with either regional lymphatic or distant metastases were associated with significantly larger tumour sizes (Tumour Stage T3 and above) (p = 0.0217, Fisher’s exact test, n=31). The median overall survival (OS) was 36.1 months (95% CI =19.8-61 months) for the entire (discovery and validation) UK cohort (Supplementary Figure 1A-C) and 64 months (95% CI = 32 months-Not Reached (NR)) for the Germany cohort (Figure 1B; Supplementary Figure 1A and B). In the UK discovery cohort, median progression-free survival (PFS) was 46.5 months (95% CI = 17.5 months-NR) for individuals with localised disease at diagnosis (Supplementary Figure 1D). Interestingly, there was a subset of six individuals in the UK cohort with a long latency (56 months - 448 months) between primary and first relapse/metastasis. A similar pattern was seen in the Germany cohort with some examples of exceptional long-term survivors (Figure 1B; Supplementary Figure 1A).

A low tumour mutational burden (TMB) (median of 1 mutation per megabase) was detected across most patient samples (Figure 1C; Supplementary Figure 1E). The exception were the samples without paired germline DNA and cell lines which despite extensive post-processing filtering still harboured substantial private single nucleotide polymorphisms. Recurrent hotspot variants were identified in *TERT*, a gene that encodes for the catalytic subunit of telomerase, (c.-124C>T or c.-146C>T), a cis-regulatory element mutation, hypermethylation of the promoter CpG island (cg11625005) and putative “enhancer hijacking” of the promoter region were identified in 12 (19.6%), 1 (1.6%), 6 (9.8%) and 1 (1.6%) patient sample/s respectively (Figure 1C). We then extended the analysis to the expansion cohort using archival FFPE samples (n=26) (Supplementary Table 2). This analysis revealed that 8/26 cases were either *TERT* promoter hypermethylated (*n*=3) or had canonical *TERT* promoter mutations (*n*=5) yielding an overall prevalence of 32.18% (*n* = 28/87) with pathogenic *TERT* promoter alterations. Of note, primary tumours with synchronous metastases were enriched in *TERT* alterations (p = 0.035, Fisher’s exact test). Six individuals demonstrated inactivating frameshift, nonsense mutations or inactivating structural variants (SVs) in *MGA*, a hitherto undescribed potential cancer driver in CCS and a known transcriptional repressor of MYC activity (Hurlin et al., 1999). Less frequent but recurrent mutations (frequency < 3 samples) were identified in other cancer genes including *TP53, NF1, EIF1AX, and MTOR* (Figure 1C; Supplementary Figure 1F). The CCS cell lines showed a similar mutational repertoire as the patient samples with 3/10 possessing *TERT* promoter mutations, 1/10 possessing a *TERT* putative promoter hijacking event and 3/10 with disruptive *MGA* structural variants. The biggest distinction was a higher frequency of hotspot *TP53* mutations in the cell lines compared to patient samples (37.5% vs <5%) likely representing an *in-vitro* adaptation bias like other cancer subtypes (Brohl et al., 2014) rather than CCS specific biology.

To gain insights into the biological processes underpinning the mutational spectra in CCS, we noted a strong correlation between the non-synonymous mutation load and overall tumour mutation burden (Pearson’s r = 0.90, P < 0.001) (Supplementary Figure 1G), consistent with proportional accumulation of mutations across the genome. This finding was corroborated by single base substitution and indel mutational signature extractions which showed dominance of clock like signatures SBS1 and SBS5 (median attribution 12.27% and 45.24%) in all samples. APOBEC signatures SBS2 and SBS13 were among other prominent signatures present at lower frequencies (median attribution 4.49% and 8.2%) in a subset of CCS samples. SBS40a (median attribution 48.6%), a signature with a currently unknown aetiological mechanism, was also found in all samples. Furthermore, clock-like indel signatures (ID1 and ID2) and signatures of unknown aetiology (ID5 and ID9) were also present in the majority of CCS samples.

### CCS harbours pervasive low-level aneuploidy

The predominant ploidy across samples was constrained at a near-diploid state (mean ploidy of 2.2N) corroborated by a pervasive CN1 (diploid) copy number signature across the cohort. However, most samples demonstrated a gain or loss of at least one entire chromosome or chromosome arm with significant variation in the fraction of genome altered (FGA), a measure of the extent of copy number alterations (CNAs), (median = 0.17, CV=0.68, p<0.05) detected between samples (Supplementary Figure 2A). Co-attribution of CN2 and CN9 signatures with CN1 further reflects an aneuploid state across the cohort (Figure 1C). CN2, a signature associated with tetraploidy showed a 100% attribution in 2 samples (CCS_22GP-T & CCS-E-P03_T) indicative of whole genome doubling (WGD) and confirmed via flow cytometry for CCS_22GP-T. Interestingly, CCS_22GP-T represented an outlier in terms of size of tumour (700 mm in maximum dimension) compared to diploid tumours. WGD is a poor prognostic feature in cancer (Bielski et al., 2018) and is a frequent occurrence in many other cancer types but has not been previously documented in CCS. To assess the frequency of WGD in CCS, we performed DNA ploidy flow cytometry analysis (methods, Supplementary Figure 2B) on tumour samples in the expansion cohort and identified an additional two primary samples with WGD, resulting in an overall incidence of 6.25% (4/64). The extent of aneuploidy and rare WGD samples seen in our CCS samples, are supported by previous karyotyping studies (Bridge et al., 1991) (Panagopoulos et al., 2002) from the pre-next generation sequencing era but have not previously been evaluated in the context of the mutational profile or prognosis. To investigate this phenomenon further, we compared our cohort of CCS cases to the NHS Genomics England 100,000 Genomes sarcoma WGS cohort of > 1000 sarcomas, comparing the fraction of genome altered (FGA) of known fusion driven sarcomas to canonically karyotypically complex sarcomas. This analysis revealed that CCS shows a significantly higher level of FGA compared to other fusion-driven sarcomas implying a higher level of genomic instability (Figure 2A; Supplementary Figure 2C). Since elevated FGA frequently co-occurs with *TP53* mutations and/or loss of heterozygosity (LOH) (Taylor et al., 2018), we assessed the correlation between them and found samples with *TP53* LOH to be significantly enriched with higher FGA (Supplementary Figure 2D) in CCS.

**Figure 2:**
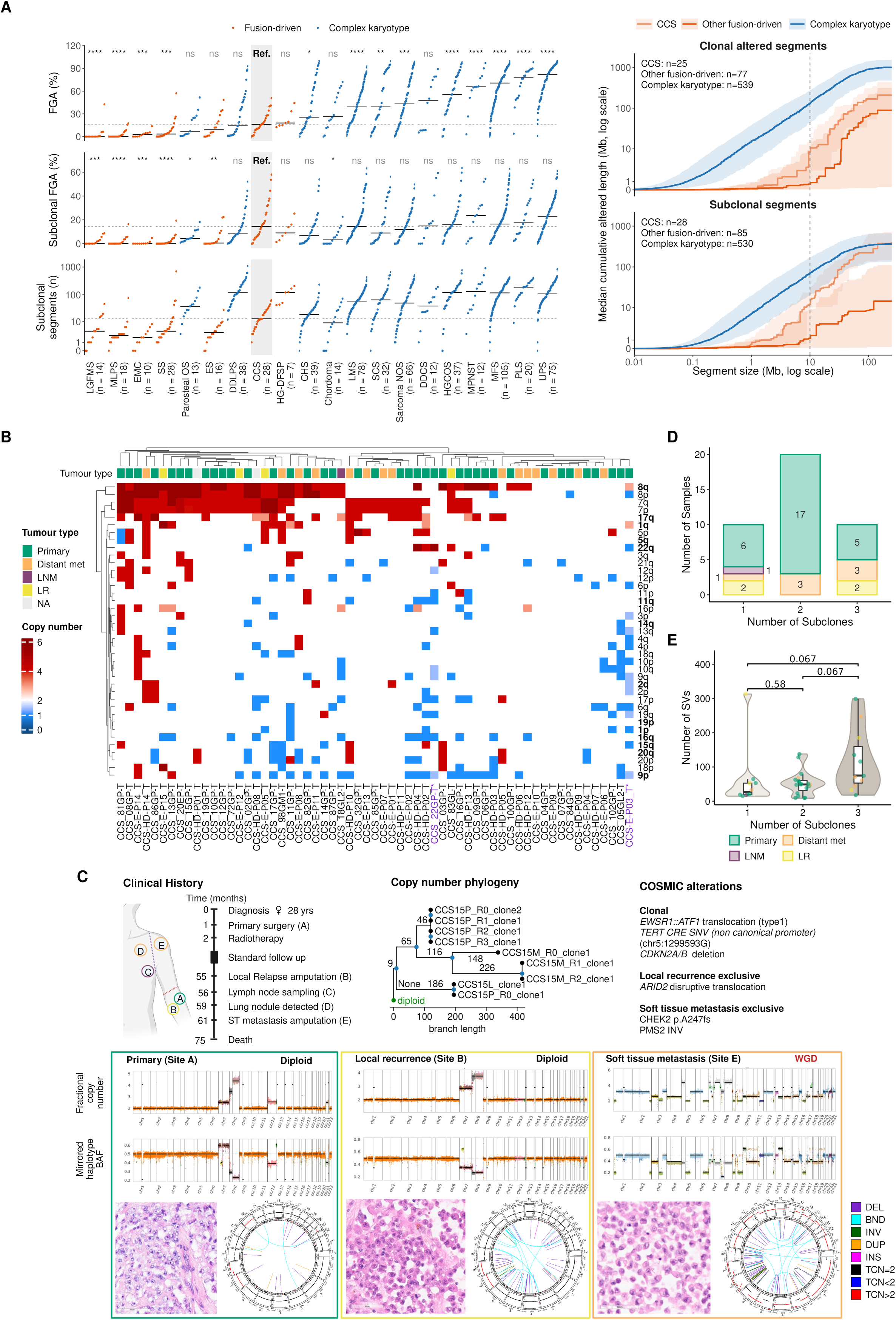
Aneuploidy is a hallmark of Clear Cell Sarcoma and drives subclonal diversification A: Left Panel: Snake plots showing (top) the fraction of the genome altered (FGA), (middle) subclonal FGA and (bottom) the number of subclonal copy-number segments for each sarcoma subtype. Each point represents one tumour; within each subtype, tumours are plotted in ascending order of their value. Subtypes are ordered by median FGA, and the number of tumours per subtype is shown in parentheses. Black horizontal lines indicate subtype medians, and the shaded grey box indicates clear cell sarcoma (CCS) serves as the reference. Points are coloured by tumour group: fusion-driven (blue) or complex karyotype (orange). Segment counts in (bottom) are shown on a log scale. Each subtype was compared with CCS using a two-sided Wilcoxon rank-sum test with Benjamini–Hochberg correction (ns, not significant; *P < 0.05; **P < 0.01; ***P < 0.001; ****P < 0.0001). Subtypes with fewer than five tumours were excluded. CCS- Clear cell sarcoma; LMS - Leiomyosarcoma ; PLS - Pleomorphic liposarcoma; MLPS - Myxoid liposarcoma; SS - Synovial sarcoma; EMC - Extraskeletal myxoid chondrosarcoma; MFS - Myxofibrosarcoma; HGCOS - High grade central osteosarcoma; CHS- Chondrosarcoma; DDCS - Dedifferentiated chondrosarcoma; LGFMS - Low grade fibromyxoid sarcoma; Parosteal OS - Parosteal osteosarcoma; UPS - Undifferentiated pleomorphic sarcoma; HG-DFSP - High grade Dermatofibrosarcoma protuberans; DDLPS - Dedifferentiated liposarcoma; MPNST - Malignant Peripheral Nerve Sheath Tumour ; ES - Ewing sarcoma; SCS - Spindle cell sarcoma A: Right Panel: Size distribution and burden of clonal and subclonal copy-number alterations in CCS compared with other fusion-driven and complex-karyotype sarcomas. Cumulative altered genome length (Mb, pseudo-log scale) plotted against segment size (Mb, log scale) for clonal (top) and subclonal (bottom) copy-number segments. Segments were derived from Battenberg allele-specific copy-number profiles and restricted to autosomes. A segment was considered altered when its allele-specific copy-number state differed from the expected normal state (1+1, or 2+2 in whole-genome-doubled tumours), including copy-neutral loss of heterozygosity. Clonal segments were those with a single state present in all tumour cells; subclonal segments were those fitted as a mixture of two states. Each tumour’s cumulative altered length was evaluated at 400 fixed segment sizes evenly spaced on a log scale from 0.01 to 250 Mb, and the median and interquartile range were calculated at each size. Lines show the median and shaded bands show the interquartile range (25th–75th percentile) across tumour subgroups at each segment size, for CCS (light blue), other fusion-driven sarcomas (dark blue) and complex-karyotype sarcomas (orange). The end point of each line reflects the median total altered length, and the position along the x-axis at which it rises reflects the sizes of the segments contributing to it. Tumours with at least one altered segment in the respective category were included; the number of tumours per group is shown in each panel. The dashed vertical line marks 10 Mb. B: Hierarchically clustered heatmap of arm-level copy-number alterations across CCS using GISTIC. Each column represents a tumour and each row a chromosome arm. Copy-number gains and losses are depicted in red and blue, respectively, with colour intensity reflecting copy-number state. Dendrograms show unsupervised hierarchical clustering of tumours and chromosome arms. Tumour type is indicated by the colour annotation bar above the heatmap (Primary, lymph node metastasis (LNM), distant metastasis, local recurrence (LR), or not available (NA)). Chromosome arms shown in bold on the right-hand axis correspond to significantly recurrent copy-number alterations identified by GISTIC analysis. Purple coloured x-axis labels denote samples with whole genome doubling (WGD). C: Clinical course, copy-number phylogeny, genomic profiles, and histological features from a patient with CCS sampled longitudinally. Left, schematic timeline showing disease course, anatomical sites sampled, and timing of interventions. Top centre, MEDICC-derived phylogeny reconstructed from whole-genome sequencing copy-number profiles of spatially and temporally distinct tumour samples. Branch lengths are proportional to copy-number evolutionary distance. Top right, selected somatic alterations identified across the tumour lineage, grouped as clonal or site-specific events. Bottom panels show representative samples from the primary tumour (site A), local recurrence (site B), and soft tissue metastasis (site E). For each specimen, haplotype-specific copy-number and mirrored B-allele frequency profiles are shown above matched haematoxylin and eosin (H&E)-stained sections and Circos plots summarising structural variants and copy-number alterations. Copy-number states are displayed as fractional copy number across the genome. Whole-genome doubling (WGD) identified in the metastatic specimen is indicated. D: Distribution of the number of subclones identified in individual tumours using DPClust. Stacked bars show the number of samples harbouring one, two, or three subclonal populations. Colours indicate tumour type (Primary, primary tumour; LR, local recurrence; LNM, lymph node metastasis; Distant met, distant metastasis) and embedded numbers denote sample counts within each category. E: Number of structural variants (SVs) per tumour sample, grouped by the number of subclones inferred by DPClust (1 subclone, *n* = 10; 2 subclones, *n* = 20; 3 subclones, *n* = 10). Violin plot shows the distribution of SV burden in each group. Boxes indicate the median (centre line) and interquartile range, with whiskers extending to 1.5× the interquartile range. Each point represents an individual sample coloured according to specimen type (Primary, primary tumour; LR, local recurrence; LNM, lymph node metastasis; Distant met, distant metastasis). P-values are from two-sided Wilcoxon rank-sum tests, adjusted for multiple comparisons with the Benjamini–Hochberg method.

While CCS showed an intermediate level of overall FGA, this measure alone cannot distinguish alterations acquired early in tumour evolution from those arising later. We, therefore, examined subclonal copy-number alterations which arise after the emergence of the most recent common ancestor, and compared them across sarcoma subtypes. CCS showed a median subclonal FGA of 14.5% which is closer to complex-karyotype sarcomas than its overall FGA would suggest, and higher than other fusion-driven subtypes (Figure 2A). However, the nature of this subclonal alteration differed markedly between CCS and complex karyotypes. CCS tumours carried a median of 20 subclonal segments, compared to multiple complex sarcomas which had a mean of per-sample median of ∼82 subclonal segments, despite a broadly comparable fraction of the genome being affected. We next asked whether the alterations in CCS resembled the fragmented pattern of complex-karyotype sarcomas. Cumulative distribution of clonal and subclonal segments (Figure 2A) confirmed that in CCS, most altered length came from segments larger than 10 Mb while in complex-karyotype sarcomas, a substantial share comes from smaller segments. Other fusion-driven sarcomas had little altered genome at any segment size. Subclonal change in CCS therefore appears to involve fewer, larger events, such as arm-level gains and losses, whereas in complex sarcomas like Undifferentiated Pleomorphic Sarcoma (UPS), it is distributed across many small segments, consistent with genome “catastrophe” processes such as chromothripsis.

Recurrent chromosomal CNAs of CCS have previously been described (Rasmussen et al., 2023) (J. Li et al., 2021) (Panza et al., 2021) with frequent gains of chromosome 1q, 7, 8 and 17q, and losses of chromosome 9 which are also seen in our cohort. Loss of chromosome arm 19p were also observed less frequently (Figure 2B). These gains and losses were broad overall, encompassing entire chromosomes or arms rather than focal genomic loci (Figure 2B; Supplementary Figure 2E). The only exception was a recurrent small deletion in chr3 encompassing the tumour suppressor and fragile site *FHIT*, which has been associated with replication stress (Waters et al., 2014) (Simón-Carrasco et al., 2024), that was observed in 8.1% (n=5) of cases. Unsupervised hierarchical clustering of allele-informed CNAs revealed two copy number clusters namely, “loss-biased” and “gain-biased” genomic phenotypes, reflecting the balance of aneuploidy within a sample shifting either towards losses and loss of heterozygosity (LOH) or gains and amplifications. Both clusters were reproducible under bootstrap resampling (mean Jaccard similarity 0.72 and 0.81), and silhouette analysis favoured k = 2 over higher partitions (mean silhouette width 0.21), indicating stable groups. Within the loss-biased cohort, disruption in chr9p (e.g. *CDKN2A/B, MTAP*) frequently co-occurred with deletions and LOH in chromosomes 10 (e.g. PTEN), 16q (e.g. *CDH1, CYLD*), 17p (e.g. *TP53, HIC1*), and 18q (e.g. *SMAD2/4, DCC*) (Figure 2B). This finding combined with gene specific LOH on a patient specific basis reveals large scale tumour-suppressor disruption, disturbing key biological pathways including the cell cycle (*CDKN2A),* DNA damage response *(TP53, CHEK2),* RAS/MAPK signalling *(NF1, SPRED2),* and chromatin regulation *(SMARCA2* and *ARID1B)*. The gain-biased samples show strong co-occurrent gains of chromosomes 8q (e.g. *MYC, RAD21*), 7q (e.g. *EGFR, MET*) and 17q (E*RBB2, BIRC5, TOP2A*) (Figure 2B). These chromosomes are commonly gained or amplified across many other cancers due to the density of key oncogenes on these loci. The gained-biased cohort also had an enrichment of chr1q gain often concurrent with pericentromeric losses on chromosome 1, another genomic fragile site often deleted during replicative stress.

Interestingly amongst the loss-biased phenotype, cases show a mirrored pattern of 17q gain with 17p loss, consistent with a possible isochromosome 17q event or other whole-arm copy-number alteration affecting chromosome 17. The gain-biased cohort shows possible evidence of isochromosome 8q development in some cases, a finding seen in published CCS karyotypes (Mitelman F, 2026) (J. Wang & Laframboise, 2019) (Morales et al., 2022) (P Dal Cin, 1999) and other sarcomas (Cortes-Ciriano et al., 2023). Besides the gain of *MYC*, chromosome 8 gains are postulated to buffer replication stress by upregulating *RAD21* in Ewing Sarcoma (Su et al., 2021) and *TMPRSS2::ERG* driven prostate cancer (Su et al., 2024), a mechanism that may be relevant in CCS given the recurrent *FHIT* deletion and pericentromeric chr1q loss observed here.

To test whether recurrent arm-level CNAs arise through whole-chromosome missegregation, we assessed co-occurrence of p and q arm alterations within each chromosome. We found significant co-segregation of both arms for gains of chromosomes 7, 5, 12 and 20 and for losses of chromosome 20, by DISCOVER (Canisius et al., 2016), a co-occurrence test that accounts for differences in overall alteration burden between tumours (q < 0.05) (Supplementary Table 3). Chromosome 7 gain (chr7 gain) was almost exclusively at whole-chromosome levels (32/34 samples in which a gain was identified; DISCOVER q = 0.02).

Chromosome 8 gain (chr8 gain) showed strong co-segregation (28 of 39 tumours), although this did not reach statistical significance (DISCOVER q = 0.08), similar to losses of chromosomes 9 (DISCOVER q = 0.09) and 10 (DISCOVER q = 0.05). In contrast, chromosomes 1, 8, 16 and 17 showed reciprocal imbalances between their two arms, however none of the association reached statistical significance. Loss of chromosome arm 1p was accompanied by gain of chromosome arm 1q in 4/5 tumours with 1p loss, gain of chromosome arm 16p by loss of chromosome arm 16q in 5/7 tumours, chromosome arm 17p loss with chromosome arm 17q gain in 5/7 tumours and chromosome arm 8p loss with chromosome arm 8q gain in 2/3 tumours, configurations consistent with potential isochromosome formation. Together these findings indicate that several recurrent aneuploidies arise through whole-chromosome missegregation, while others may reflect centromere-associated arm-level rearrangements.

The recurrence of chr7, chr8 and chromosome 17q (chr17q) gains across CCS prompted us to investigate their mutational timing relative to the founding fusion event. Using mutation content within gained segments to infer temporal hierarchy (Baker et al., 2024), chr8 gain was predominantly a truncal event and typically preceded the other recurrent gains, though its timing varied across the cases (Supplementary Figure 2F). Interestingly, the early initial gains on chr8 were associated with shorter survival than late initial gains in chr8 (median 1.1 versus 4.2 years; HR 6.37, 95% CI 1.97-23.3, p = 0.002; q = 0.013) an association that persisted even when using the highest confidence mutation-time calls (Supplementary Figure 2F). Chr7 initial gains showed no conserved timing across cases with clonal gains split evenly between early and late. In contrast, chr17q gain was consistently acquired late in mutation time, with many instances being subclonal consistent with ongoing chromosomal instability rather than a fixed early event. Timing of the chr7 and 17q gains was not associated with survival. The early truncal positioning of chr8 gain carrying *MYC* and *RAD21* among its oncogenic content suggests it is incorporated into the founding genomic programme of CCS. The late and subclonal nature of chr17q gain is notable given its association with adverse clinical outcome, consistent with a model in which 17q amplification encompassing *ERBB2, BIRC5* and *TOP2A,* genes with prognostic and therapeutic relevance in cancer, confers selective advantage during progression rather than at tumour initiation.

### Aneuploidy is associated with subclonal diversification

Despite sharing a common oncogenic fusion, CCS tumours exhibit striking variability in clinical behaviour and therapeutic resistance. We hypothesised that recurrent aneuploidies may drive subclonal diversification in CCS, with distinct aneuploid clones expanding under positive selection to generate the inter-tumour heterogeneity that underlies clinical variability.

CCS15, a patient with an exceptional long progression-free clinical course illustrates this process. This patient presented with a surgically resected and irradiated primary tumour, developed local recurrence (LR) after a prolonged disease-free interval, and subsequently progressed rapidly to lymph node and systemic metastases within two years of LR (Figure 2C). Genomic profiling across all three timepoints revealed diploid copy number profiles in both the primary and local recurrence, while the distant metastasis harboured WGD, a rapid transition that raised the possibility whether the WGD population was pre-existing at a subclonal frequency in the primary tumour.

To address this, we performed multi-region sequencing of four spatially distinct regions from the primary tumour (CCS15P_R0, CCS15P_R1, CCS15P_R2, and CCS15P_R3) alongside the LR and distant metastasis. Chr8 gains were shared across all primary tumour regions, confirming their truncal origin and early acquisition (Supplementary Figure 2G). In contrast, chr12 gain was clonal in three regions but subclonal or absent in the fourth, and gains in chr7 and chr17q were spatially confined to individual regions, consistent with a branched mode of evolution in which distinct subclonal populations occupy spatially restricted niches. Both the local recurrence and distant metastasis harboured clonal chr7 and chr8 gains with subclonal chr17q gain. Notably, the chr7 gain in the primary tumour occurred on the alternate haplotype relative to the local recurrence and metastasis, a pattern of convergent evolution implying strong positive selection for chr7 dosage independent of the specific haplotype gained. Copy number phylogenetic reconstruction confirmed independent acquisition of chr7 gain across the primary, local recurrence and metastatic lesions, and indicated early metastatic seeding from the primary tumour (Figure 2C; Supplementary Figure 2G). A similar pattern was observed in patient CCS08, in which gains of chr7q, and chr8 gain were acquired independently by primary and metastatic tumours as evidenced by the opposite haplotype (Supplementary Figure 2H), indicating convergent evolution of these CNAs.

To determine whether subclonal diversification is a general feature of CCS rather than specific to this case, we performed subclonal reconstruction across all primary tumours using bulk WGS. Most cases harboured between two and four distinct genetic subclones, with the majority demonstrating at least two (Figure 2D). Samples with greater subclonal complexity showed higher structural variant burden, although this association did not remain significant after correction for multiple testing (Figure 2E). Similarly, the association between subclonal complexity and FGA did not reach significance at all (Supplementary Figure 2I). Together, these findings establish that CCS is not a clonally homogeneous disease, and that structural genomic complexity, rather than simple copy number burden is the primary correlate of subclonal diversification.

### Complex structural events are a prominent feature of the SV landscape

Whole-genome sequencing revealed a median of 51 SVs per tumour (range 9–313) (Supplementary Figure 3A), with deletions the most frequent class (33.6%), followed by inversions (25.5%), duplications (18.8%) and breakend junctions representing complex or interchromosomal rearrangements (21.3%). Beyond the oncogenic fusion, no recurrent SVs were identified across the cohort, though a subset of cases harboured markedly elevated SV burden (>100 SVs; n=8). Strikingly, complex structural events including chromoplexy, chromothripsis and chained open-loop rearrangements were present in 50% of cases (Figure 3A-C; Supplementary Figure 3B-D), a finding that differs from the prevailing characterisation of CCS as a genomically simple disease.

**Figure 3:**
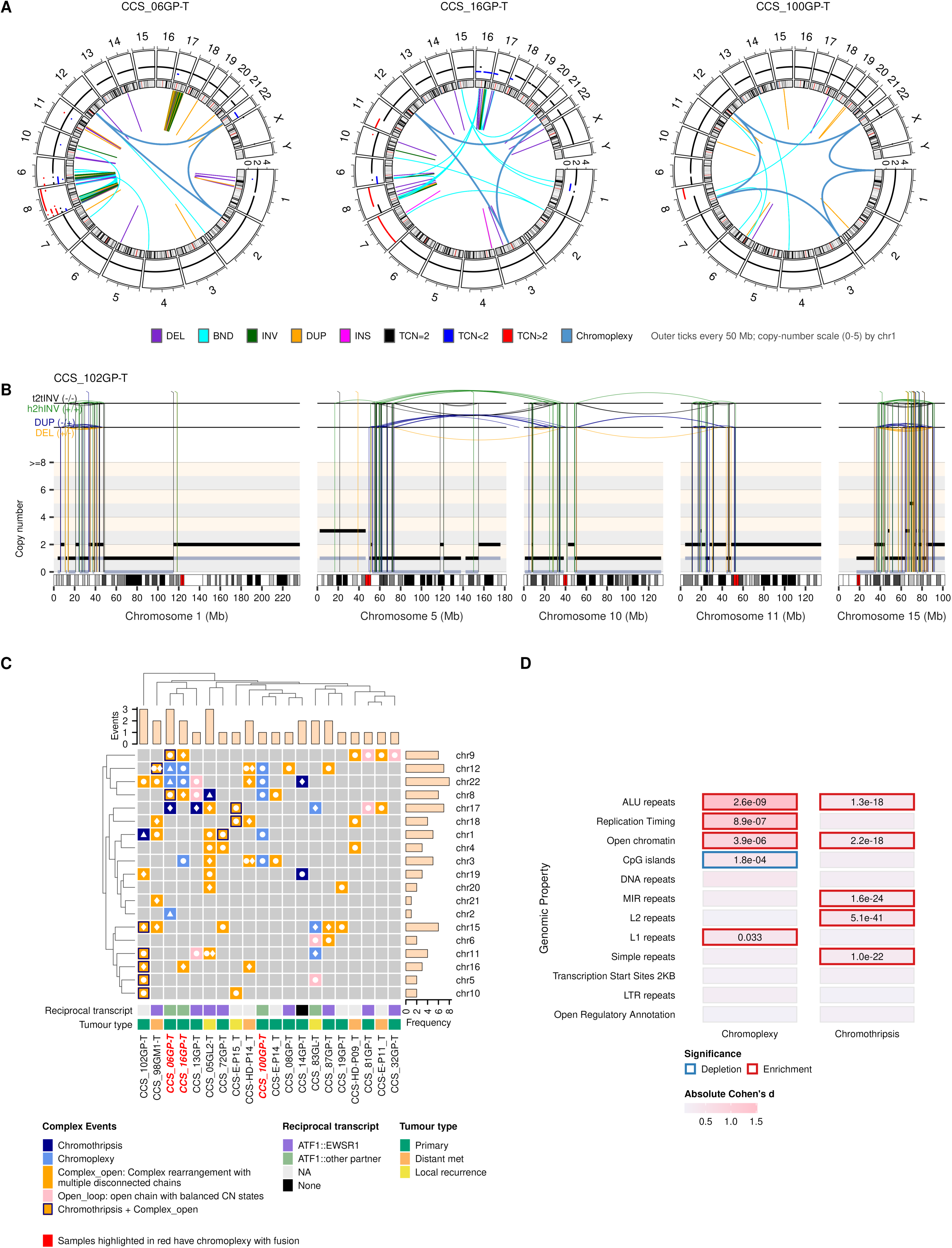
Chromothripsis and chromoplexy shape the structural variant landscape of a subset of CCS A: Circos plots depicting whole-genome sequencing-derived structural variants and copy-number alterations. Chromosomes are arranged sequentially around the outer ring. The outer track shows total copy number, with coloured segments indicating copy-number gains and losses. Inner links represent structural variants, coloured according to rearrangement class (deletions, breakends, inversions, and duplications). Blue arcs highlight structural variant chains assigned to chromoplexy events. B: Copy-number and structural variant profiles for selected chromosomes from CCS102 demonstrating chromothripsis. Chromosomes are displayed according to genomic position (Mb). Total copy number is represented by black horizontal segments. Structural variants are shown as arcs connecting breakpoint pairs and coloured by rearrangement type (deletion, duplication, head to head inversions, and tail to tail inversions). Cytobands are shown beneath each chromosome, with centromeres highlighted in red. The figure illustrates extensive clustering of breakpoints and copy-number oscillations across multiple chromosomes. C: Heatmap showing chromosomes involved in complex structural variant events across the cohort. Rows represent chromosomes and columns represent individual tumour samples. Event classes include chromothripsis, chromoplexy, complex open-chain rearrangements, open-loop rearrangements, and co-occurring chromothripsis with complex open-chain rearrangements. For chained rearrangements, identical symbols (e.g., circles, diamonds, triangles) denote chromosomes participating in the same event, whereas different symbols within a sample indicate independent complex events. Samples and chromosomes were hierarchically clustered based on event profiles. Top and right bar plots indicate the number of complex events per sample and the frequency of chromosome involvement, respectively. Annotation tracks denote reciprocal fusion transcript status and tumour type. Samples highlighted in red harbour chromoplexy events linked to the EWSR1::ATF1 fusion. D: Association of chromoplexy and chromothripsis breakpoints with genomic features. Heatmap colours denote absolute effect size (Cohen’s d), and coloured outlines indicate significant enrichment (red) or depletion (blue); corresponding P values are shown.

Most tumours demonstrated a conventional balanced translocation generating the *EWSR1::ATF1* or *EWSR1::CREB1* fusion, with a detectable reciprocal in-frame *ATF1::EWSR1* transcript in all such cases. However, 3/65 tumours harboured chromoplexy-mediated fusions, including one case (CCS_100GP-T) in which five chromosomes participated in a single chromoplectic event (Figure 3A). Chromoplexy-mediated fusions were not accompanied by reciprocal transcripts but instead produced ATF1 fusions with alternative partner genes arising from the chromoplectic loop for example *ATF1::NXPE3* in CCS_16GP-T. Notably, alternative ATF1 fusion transcripts cannot be used as a reliable proxy for chromoplexy, as cases with conventional balanced translocations also expressed non-canonical ATF1 transcripts arising from concurrent complex SVs including large deletions and inversions at the fusion locus. Compared to Ewing sarcoma, where chromoplexy-mediated fusions occur at higher frequency (Anderson et al., 2018), CCS shows a lower overall rate of fusion complexity, higher instances of reciprocal transcript expression and distinctive ATF1 transcriptional consequences when chromoplexy does occur.

Unsupervised hierarchical clustering of complex variant profiles separated tumours into two groups reflecting differences in the frequency, chromosomal distribution and composition of complex rearrangement classes (Figure 3C). High-complexity tumours (Cluster A) were characterised by recurrent chromothripsis, chromoplexy, and compound chromothripsis-open loop events spanning multiple chromosomes. Low-complexity tumours (Cluster B) showed sparse SV profiles typically limited to one or two isolated events confined to individual chromosomes. Co-occurrence of rearrangements across the same chromosomes in Cluster A samples, particularly chromosomes 12 and 22, encompassing *ATF1* and *EWSR1*, suggests preferential co-rearrangement of these loci in tumours with high structural complexity.

To characterise the genomic context in which these complex events arise and the signs of their repair mechanisms, we performed enrichment analysis of genome topography at chromoplectic and chromothriptic breakpoints. Chromoplectic breakpoints were significantly enriched at ALU repeats, L1 repeats, open chromatin regions and early-replicating domains (Figure 3D; Supplementary Figure 3E), consistent with replication fork collapse at transcriptionally active origins triggering template switching between proximal repetitive elements during replication stress, the likely mechanistic basis for the multi-chromosomal chained rearrangements characteristic of chromoplexy (Carvalho & Lupski, 2016) (Baca et al., 2013). Chromothriptic breakpoints were instead enriched at ALU, MIR, L2 and simple repeats (Figure 3D), implicating microhomology-mediated end joining as the predominant reassembly mechanism following chromosomal shattering, mechanistically distinct from the replication-coupled process inferred for chromoplexy (Cortés-Ciriano et al., 2020) (Umbreit et al., 2020). Chromoplectic breakpoints were conversely depleted at CpG islands.

Consistent with the generation of abundant single-stranded DNA during chromothriptic remodelling, chromothriptic samples showed significant enrichment of the APOBEC-associated signature SBS13 (Fisher’s exact test p=0.02; OR=8.8, 95% CI 1.08–86.9). This finding indicates that chromothripsis in CCS is not solely a source of structural rearrangement but also a trigger for localised APOBEC-mediated mutagenesis, potentially increasing sequence-level genomic diversity beyond the structural changes themselves and contributing to the subclonal diversification documented in the preceding section.

### Transcriptional consequences of aneuploidy

Having established the pervasive copy number landscape of CCS including gain-biased and loss-biased genomic phenotypes and recurrent focal events at clinically relevant loci we next asked how these specific aneuploidies shape gene expression and tumour behaviour. PCA of high-depth bulk mRNA sequencing data from the UK cohort (28/32 primary tumours) identified two dominant axes of transcriptional variation (Figure 4A). PC1 (15.1% variance) segregated samples by tumour purity and captured a continuum between immune-enriched states marked by interferon-γ response and tumour-intrinsic inflammatory programmes enriched for TNFα/NF-κB and IL-6/JAK-STAT3 signalling. PC2 (11.8% variance) was significantly correlated with FGA and was enriched for G2M checkpoint, E2F target, mitotic spindle and glycolysis gene sets, reflecting a highly proliferative and metabolically active transcriptional state. High-FGA tumours were preferentially distributed toward extreme PC2 values, indicating that aneuploidy burden in CCS promotes tumour-intrinsic proliferative and Warburg metabolic programmes rather than producing broad unbuffered expression changes across the transcriptome. Individuals presenting with synchronous metastasis or short progression-free survival clustered within regions of PC space characterised by high proliferative activity and high tumour purity, consistent with an immune-excluded, rapidly proliferating tumour state (Supplementary Figure 4A).

**Figure 4:**
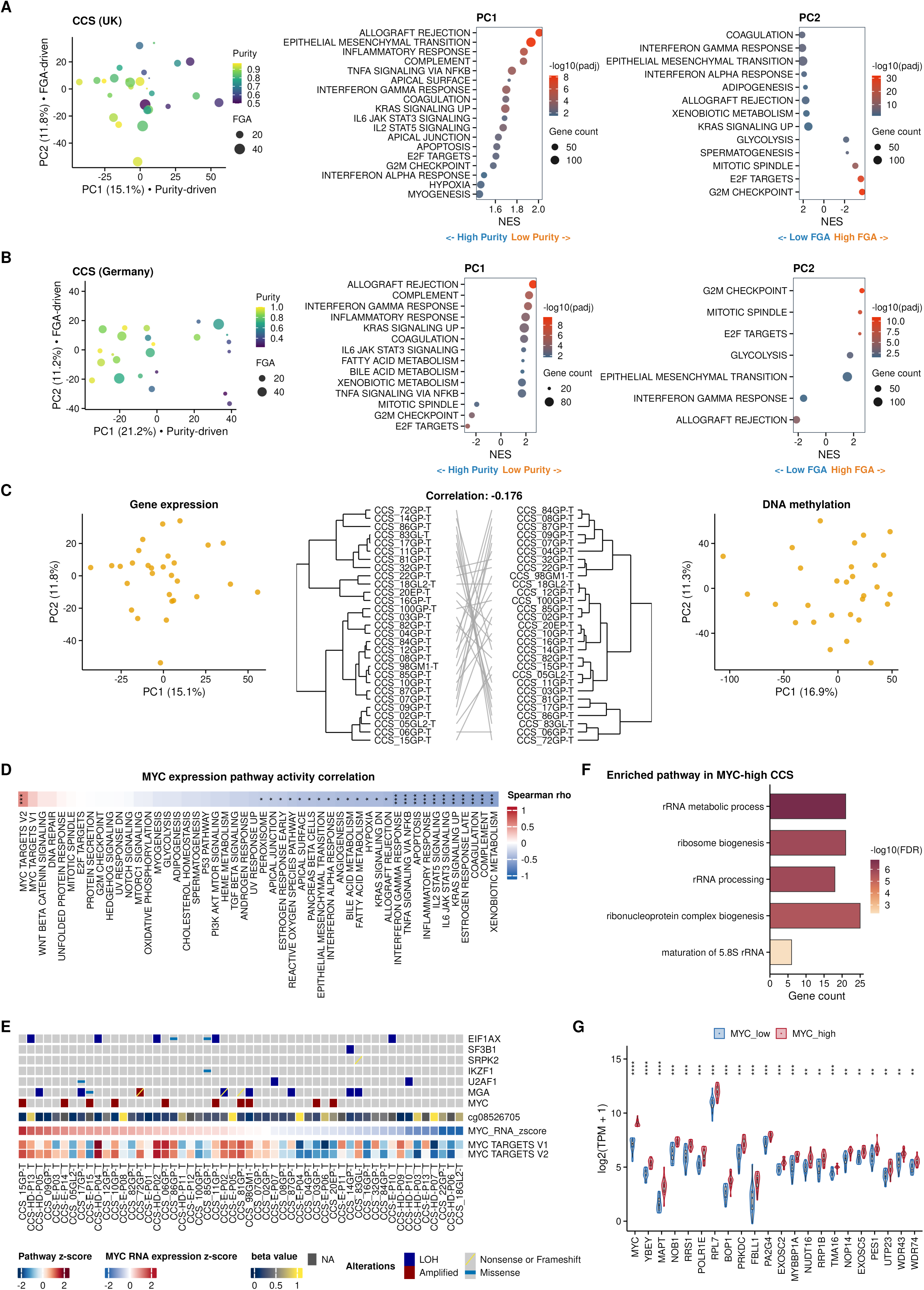
Transcriptional consequences of Aneuploidy A: Left panel: Principal component analysis (PCA) of bulk RNA-sequencing data (UK cohort). Each point represents a tumour sample. Point colour denotes tumour purity and point size denotes fraction of genome altered (FGA). PC1 and PC2 explain 15.1% and 11.8% of transcriptional variance, respectively. PC1 segregated samples according to tumour purity (Spearman’s rho= -0.616, *p*-value= 0.00037) while PC2 was significantly correlated with FGA (Spearman’s rho= - 0.477, *p*-value= 0.0095). Middle and right panel: Gene set enrichment analysis (GSEA) of genes contributing to PC1 and PC2. Normalized enrichment scores (NES) are shown on the x-axis. Dot size corresponds to gene set size and colour indicates significance (-log10 adjusted P value). Positive PC1 scores were associated with low-purity samples, there were no pathways enriched in negative PC1 scores. Negative PC2 scores were associated with high-FGA tumours, whereas positive PC2 scores were associated with low-FGA tumours. B: Left panel: Principal component analysis (PCA) of bulk RNA-sequencing data (Germany cohort). Each point represents a tumour sample. Point colour denotes tumour purity and point size denotes fraction of genome altered (FGA). PC1 and PC2 explain 21.2% and 11.2% of transcriptional variance, respectively. PC1 was significantly correlated with tumour purity (Spearman’s rho= -0.73, p-value= 4.386e-05) while PC2 showed a weak non-significant correlation with FGA (Spearman’s rho= 0.131, *p*-value>0.05). Middle and right panel: Gene set enrichment analysis (GSEA) of genes contributing to PC1 and PC2. Normalized enrichment scores (NES) are shown on the x-axis. Dot size corresponds to gene set size and colour indicates significance (-log10 adjusted P value). Positive PC1 scores were associated with low-purity samples, whereas negative PC1 scores were associated with high-purity samples. Positive PC2 scores were associated with high-FGA tumours, whereas negative PC2 scores were associated with low-FGA tumours. C: Left panel: Principal component analysis (PCA) of bulk RNA-sequencing data (UK cohort). Each point represents a tumour sample. Middle panel: Correlation between global transcriptomic and DNA methylomic profiles. Tanglegram comparing hierarchical clustering dendrograms of bulk RNA-seq (left) and DNA methylation (right) profiles across matching samples. Connecting lines link identical samples across both omics layers Right panel: Principal component analysis (PCA) of DNA methylation data (UK cohort). Each point represents a tumour sample. D: Correlation analysis between MYC expression and Hallmark gene set activity scores across the batch-corrected samples. Colour denotes the correlation coefficient (ρ), and asterisks indicate significance following multiple-testing correction. Pathways are ordered according to correlation strength. HALLMARK_MYC_TARGETS_V2 showed the strongest positive correlation with MYC expression. E: Integrated heatmap of recurrent genomic alterations, methylation, MYC expression, and Hallmark MYC target signature activity across the cohort. Samples are ordered according to MYC pathway activity. Genomic alterations are annotated by event class as indicated. F: Gene ontology analysis of transcripts increased in MYC-high clear cell sarcomas. Bar length represents gene count and colour indicates significance (-log10 FDR). Selected significantly enriched biological processes are shown. G: Violin plots show expression of selected genes associated with ribosome biogenesis and rRNA processing in MYC-high and MYC-low CCS. Expression values are shown as log₂(TPM + 1). Boxes indicate the median (centre line) and interquartile range, with whiskers extending to 1.5× the interquartile range. P values were calculated using two-sided Wilcoxon rank-sum test: **P < 0.01, ***P < 0.001, ****P < 0.0001.

These findings were substantially replicated in the Germany cohort, where PC1 (21.2% variance) again captured immune-enriched versus tumour-intrinsic inflammatory variation correlated with tumour purity, and PC2 (11.2% variance) was enriched for the same proliferative and metabolic gene sets (Figure 4B). In this cohort, PC2 did not reach significance for correlation with global FGA, but genes contributing to PC2 were significantly enriched within recurrent GISTIC-defined focal copy number events, specifically gains at 20q13, 17q25 and 22q13 and losses at 14q32 and 22q11, harbouring cell-cycle and mitotic regulators including *AURKA* and *BIRC5* (Supplementary Figure 4B). This indicates that the proliferative transcriptional programme in CCS is driven by specific focal copy number events rather than global aneuploidy burden, explaining the decoupling of PC2 from FGA in this cohort. Analysis of the relationship between bulk transcriptomic profiles and genome-wide DNA methylation patterns revealed a weak inverse correlation (Figure 4C), consistent with methylation-mediated silencing contributing to the transcriptional landscape but suggesting that copy number-driven expression changes predominate over epigenetic reprogramming at the cohort level.

Together, these analyses reveal that aneuploidy in CCS shapes tumour behaviour through two separate transcriptional mechanisms: a global aneuploidy burden that promotes proliferative and glycolytic reprogramming, and specific focal copy number gains that amplify discrete oncogenic programmes including mitotic dysregulation through AURKA and BIRC5. Both programmes are associated with aggressive clinical behaviour and operate in an immune-excluded context, providing a transcriptional basis for the clinical heterogeneity that distinguishes indolent from rapidly progressive disease.

The recurrent co-occurrence of chr8 gain encompassing *MYC* and *MGA* inactivation across CCS raises the possibility of cooperative deregulation of MYC signalling, in which increased MYC dosage is further amplified by loss of MGA-mediated transcriptional repression.

Consistent with this, HALLMARK_MYC_TARGETS_V2 emerged as the top pathway positively associated with MYC mRNA expression across the cohort (Figure 4D). Tumours with lower MYC activity showed relative enrichment of coagulation, fatty acid metabolism and angiogenesis pathways, indicating that CCS partitions into transcriptional states defined either by high biosynthetic MYC activity or by stromal and metabolic programmes a distinction with potential implications for therapeutic targeting.

To understand what drives inter-tumoural differences in MYC pathway activation, we integrated genomic and epigenomic data with transcriptomic profiles. MYC RNA expression, rather than *MYC* copy number gain, was the primary correlate of MYC pathway enrichment (Figure 4E), indicating that transcriptional regulation rather than genomic amplification is the dominant determinant of MYC output in CCS. Hypermethylation at a *MYC* regulatory hotspot (EPIC V2 probe cg08526705) correlated with lower *MYC* expression and reduced pathway activity, consistent with methylation-mediated silencing at regulatory elements modulating *MYC* transcriptional output (Barry et al., 2021) an association that was most consistent in chr8-neutral cases, where copy number amplification does not override epigenetic silencing.

Network analysis of the MYC Targets V2 gene set identified a highly interconnected module enriched for rRNA processing and ribosome biogenesis genes as the central hub of MYC-associated transcriptional activity in CCS (Figure 4F). The hub spanned multiple stages of ribosome production from early small subunit processome components through large subunit biogenesis factors to nucleolar structural regulators, suggesting that *MYC*-high CCS enforces not only increased translational capacity but sustained commitment to pre-rRNA processing, subunit maturation and nucleolar integrity. Stratification of tumours into *MYC*-high and *MYC*-low groups by expression quartile confirmed significant differential expression of rRNA processing hub genes, with the strongest differences observed for *WDR43*, *NOLC1* and *NOP16* (Figure 4G), factors occupying complementary positions within the ribosome biogenesis pathway at the levels of early pre-rRNA processing, rRNA production and nucleolar organisation, respectively (Wada et al., 2014) (C. Zhang et al., 2014)(Lafita-Navarro et al., 2018)(Khan et al., 2025).

To determine whether this relationship extends to tractable experimental systems, we profiled CCS cell lines and found that MYC pathway activity and elevated expression of the rRNA processing hub genes including *WDR43*, *NOLC1* and *NOP16* were consistently reproduced across models (Supplementary Figure 4C). These data establish CCS cell lines as relevant systems for interrogating MYC-linked nucleolar dependencies and nominate RNA Pol I-dependent rRNA transcription as a candidate therapeutic vulnerability in MYC-high CCS (Drygin et al., 2011).

### Immune Landscape, HLA Diversity and T-cell Receptor Repertoire

Immune-related transcriptional programmes emerged as a primary axis of inter-tumour variation in CCS (Figure 4A–B), implicating the tumour microenvironment as a potential determinant of the clinical heterogeneity that defines this disease. To characterise the TME systematically, we performed multi-modal immune deconvolution using both DNA methylation and bulk RNA-seq data, obtaining concordant estimates of immune infiltration across modalities (Supplementary Figure 5A–B). The results were unambiguous in that the vast majority of CCS tumours were profoundly immune depleted. Direct comparison with melanoma and undifferentiated pleomorphic sarcoma (UPS), two tumour types with established immunogenicity and clinical responses to checkpoint blockade (Hugo et al., 2016) (Steele et al., 2019) (Keung et al., 2020), confirmed that CCS occupies the extreme low end (immune-desert or excluded) of the sarcoma immune spectrum, with ESTIMATE immune scores significantly lower than both comparators across both cohorts (Figure 5A). A small subset of cases, CCS_16GP-T, CCS_20EP-T, CCS_22GP-T, CCS_32GP-T, CCS_81GP-T and CCS_84GP-T showed relatively higher immune infiltration and are discussed further below (Supplementary Figure 5A).

**Figure 5:**
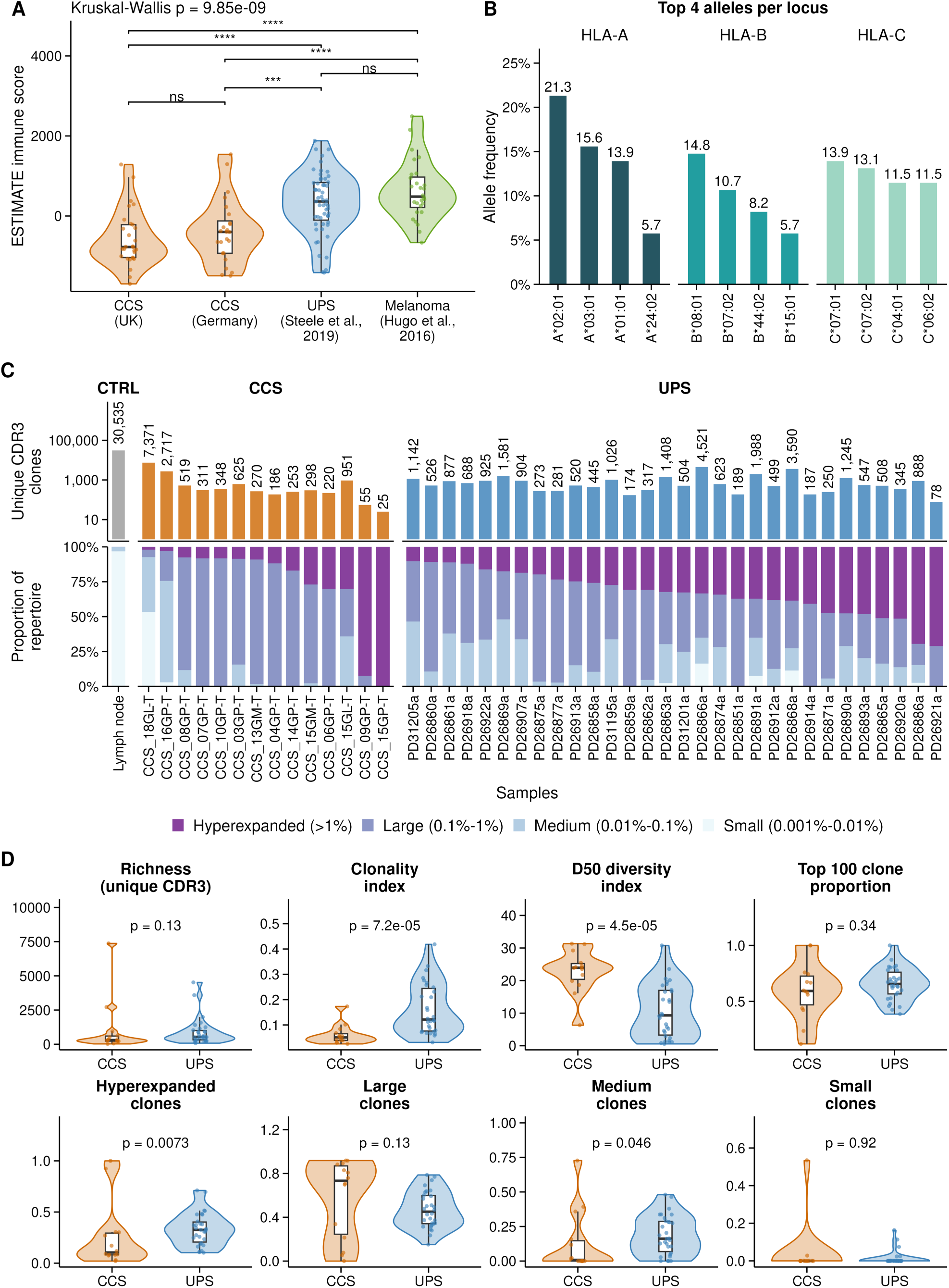
Immune Landscape, HLA Diversity and T-cell Receptor Repertoire in CCS A: Violin plots compare ESTIMATE immune scores among the UK CCS cohort, Germany CCS cohort, undifferentiated pleomorphic sarcoma (UPS), and melanoma. Boxes indicate the median and interquartile range, and points represent individual samples. Overall differences were assessed using a Kruskal-Wallis test, with pairwise comparisons shown. Statistical significance is indicated as shown (ns = not significant). B: Distribution of HLA-A, HLA-B, and HLA-C alleles across the cohort. Plot show the most frequent alleles at each locus. Frequencies are expressed as percentages. C: T cell receptor (TCR) repertoire analysis. Upper panel shows the number of unique CDR3 clonotypes identified in each sample. Lower panel shows the proportional distribution of TCR clonotypes stratified by clonal expansion category: hyperexpanded (>1% of repertoire), large (0.1%-1%), medium (0.01%-0.1%), and small (0.001%-0.01%). A reactive lymph node is included as a reference control. Samples are grouped by tumour type, with CCS shown in orange and UPS shown in blue. D: Violin plots compare TCR repertoire metrics between CCS (orange) and UPS (blue) tumours. The metrics shown are: repertoire richness (number of unique CDR3 sequences); clonality index (1 − normalised Shannon entropy); D50 diversity index (the percentage of unique clonotypes that make up 50% of the total repertoire); the proportion of the repertoire occupied by the 100 most abundant clones; and the fraction of the repertoire occupied by hyperexpanded, large, medium and small clones. Each dot represents one tumour, and embedded boxplots show the median and interquartile range. P values were calculated using two-sided Wilcoxon rank-sum tests.

Total neoantigen burden was strikingly low across the cohort (range 1-10 predicted neoantigens per sample), well below that reported for immunotherapy-responsive tumours such as melanoma (Snyder et al., 2014) and consistent with the observed low mutational burden in CCS, providing a mechanistic basis for the failure to generate tumour-reactive T-cell responses. Samples with low immune infiltration predominantly also had low neoantigen burden; notably, CCS_16GP-T, one of the cases with relatively higher immune infiltration, also displayed low neoantigen burden, suggesting that immune recruitment in this case may be driven by mechanisms independent of neoantigen load (Supplementary Figure 5C).

Critically, immune exclusion across the cohort is not explained by defects in the antigen presentation machinery: no somatic mutations or CNAs were identified in any HLA gene across all samples, and B2M was similarly intact. Tumour cells therefore retain the molecular capacity to present antigens but fail to generate the antigenic substrate necessary to trigger a productive adaptive immune response, a distinction with direct therapeutic implications, as it identifies antigen scarcity rather than antigen presentation failure as the primary barrier to immune engagement in CCS.

High-resolution germline HLA genotyping revealed that HLA-A*02:01 was the most prevalent allele in the cohort (21.3%), followed by HLA-A*03:01 (15.6%), HLA-B*08:01 (14.8%), HLA-A*01:01 (13.9%) and HLA-C*07:01 (13.9%), a distribution consistent with predominantly Northwestern European ancestry (Figure 5B; Supplementary Figure 5D). Germline homozygosity at one or more HLA-I loci was identified in 14/61 cases (22.9%) and was unexpectedly enriched among cases with relatively higher immune infiltration (p=0.045, Fisher’s exact test). No somatic mutations in HLA-A, HLA-B, HLA-C or B2M were identified, confirming that the infiltration differences are not attributable to somatic antigen presentation defects. Somatic HLA predicted loss of function was identified in only one case CCS-E-P03 indicating that homozygosity in the remainder reflects a germline diversity constraint rather than tumour-intrinsic immune escape.

TCR sequencing revealed a globally low unique CDR3 count across the CCS cohort, reflecting restricted T-cell diversity consistent with an immune-desert microenvironment (Figure 5C). Despite this low clonal richness, medium and large clones dominated the T-cell population, accounting for more than 60% of the T-cell repertoire in most cases, with hyperexpanded clones present in smaller but variable proportions. Formal comparison with UPS confirmed that CCS shows significantly lower TCR clonality, a significantly higher D50 diversity index, and a significantly lower proportion of hyperexpanded clones (Figure 5D), consistent with the absence of focused antigen-driven T-cell expansion. The sparse neoantigen profile showed a weak inverse correlation with TCR clonality, confirming that neoantigen load is insufficient to drive clonal expansion in most CCS tumours (Supplementary Figure 5C).

Against this broadly immune-desert background, two distinct outlier immune patterns were apparent in the TCR data (Figure 5C). CCS_16GP-T and CCS_18GL1-T showed high unique CDR3 counts - 2,717 and 7,371 respectively - with predominance of medium and small clones and the lowest fraction of hyperexpanded clones in the cohort (approximately 3%), suggesting recent T-cell recruitment without clonal focusing and a pattern consistent with non-specific or bystander immune activation. In contrast, CCS_09GP-T and CCS_15GP-T showed the opposite pattern: very low CDR3 diversity but a very high proportion of hyperexpanded clones exceeding 75% of the T-cell repertoire, consistent with oligoclonal antigen-driven expansion in an otherwise restricted immune context. These two patterns may reflect distinct modes of partial immune engagement in a tumour type that broadly resists productive immunity, and the oligoclonal subset in particular represents a candidate population for prospective evaluation of immunotherapeutic interventions.

The most informative immune trajectory in the cohort came from longitudinal TCR profiling of CCS15, the same patient whose multi-region genomic analysis revealed spatially restricted subclonal aneuploidies and the emergence of a WGD clone specifically at the metastatic site following a prolonged disease-free interval. TCR sequencing across the primary tumour, local recurrence and distant metastasis revealed a striking parallel evolution of the immune landscape that mirrored the genomic trajectory. T-cell diversity was lowest at the primary tumour, rose substantially at local recurrence where a unique CDR3 count of approximately 900 was observed and then collapsed at the distant metastatic site. Local recurrence in this case developed after a prolonged disease-free interval following surgical resection and post-operative radiotherapy of the primary tumour, and the restoration of T-cell diversity at this timepoint suggests that removal of the primary immune-suppressive tumour mass, combined with radiotherapy-associated immunogenic cell death, transiently restored immune surveillance.

### Genetic and non-genetic determinants of poor survival

CCS lacks validated prognostic biomarkers for routine clinical use (WHO Classification of Tumours Editorial Board, 2026). FNCLCC histological grading, the standard risk stratification framework applied across soft tissue sarcomas, has not demonstrated prognostic value in CCS, which are regarded as high-grade by definition. This over-simplification leaves clinicians reliant on stage and tumour size alone, with no means of further stratifying risk among individuals with localised disease. We therefore examined whether the genomic features characterised in this study could provide clinically useful prognostic information. Overall survival varied substantially across the cohort despite shared histological diagnosis (Figure 1B; Supplementary Figure 1A-C), and we systematically evaluated candidate prognostic parameters in primary tumour samples only (Supplementary Table 4; Supplementary Figure 6A-C) through univariate log-rank test and Cox regression analysis. Baseline metastatic status was the strongest single predictor of overall survival, with systemic spread conferring worse prognosis than lymphatic spread (Figure 6A), consistent with prior reports in CCS and other soft tissue sarcomas (H. Y. Wang et al., 2025) (H. Liu et al., 2022). Additional parameters significantly associated with poor outcome on univariate analysis were primary tumour size, chr17q gain, deletion of 9p21.3, and presence of *TERT* promoter mutation or *TERT* promoter hypermethylation (pTERT; Figure 6B-E). Chr17q gain was validated by fluorescence in-situ hybridisation (FISH) or EPICv2 microarray profiling in cases for which sequencing data were unavailable (expansion cohort) (Figure 6D; Supplementary Table 5). Similarly, to increase statistical power, we performed univariate analysis of primary tumour size and pTERT in the combined discovery and expansion cohort where data was available. CCS cases were additionally stratified by CINSARC classification, a transcriptional chromosomal complexity index developed for risk stratification across soft tissue sarcomas (Chibon et al., 2010), using the original scoring strategy. CINSARC high-risk cases had significantly worse overall survival than low-risk cases (Figure 6F).

**Figure 6:**
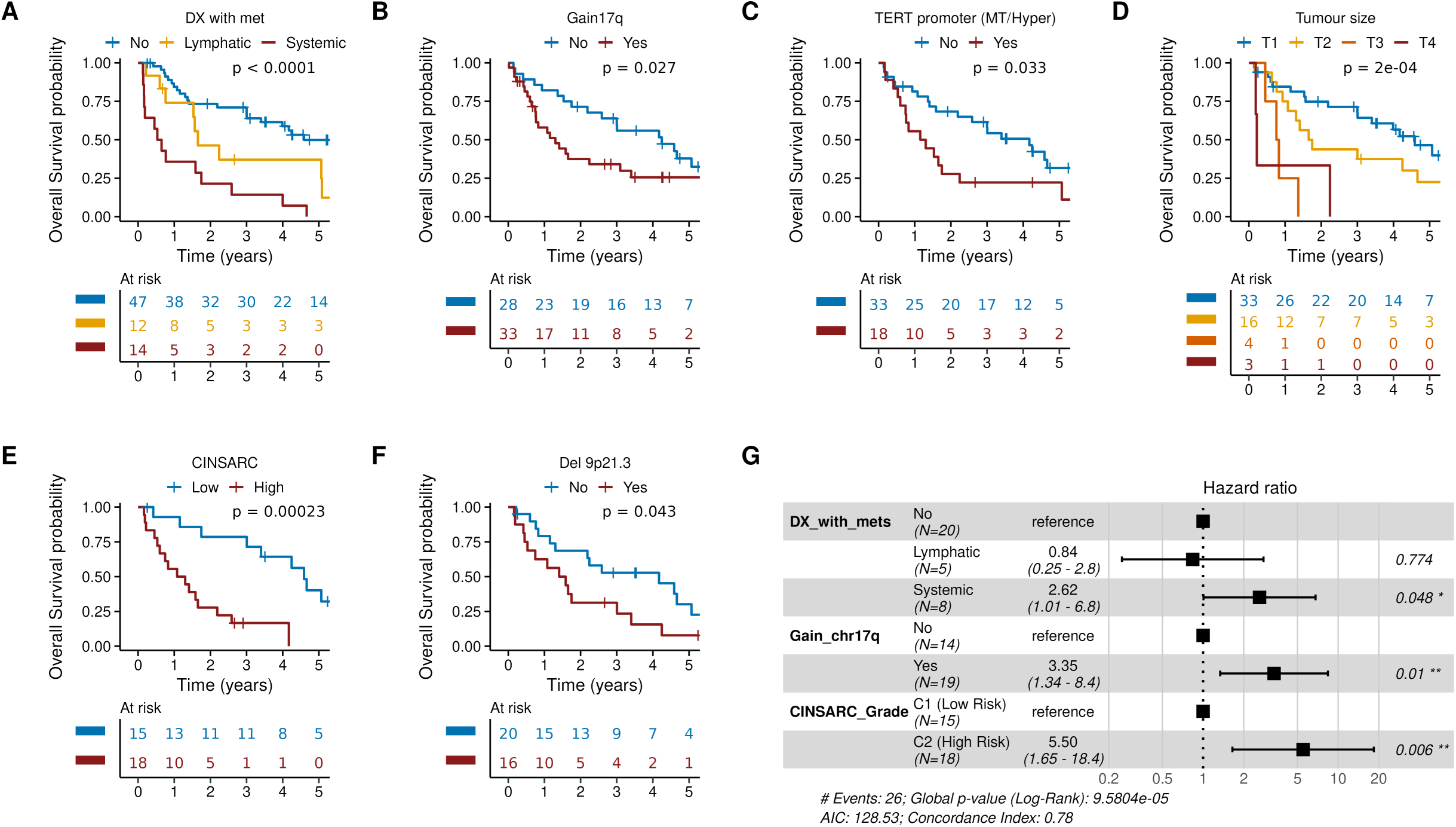
Genetic and non-genetic determinants of survival in CCS A-F: Kaplan-Meier overall survival curves stratified by clinical and genomic variables. Survival differences were assessed using two-sided log-rank tests. Tick marks indicate censored observations, and numbers at risk are shown below each plot. Kaplan–Meier curves are shown up to 5 years; log-rank P-values were computed over the entire follow-up period. Variables examined include (A) metastatic status at diagnosis, (B) chromosome 17q gain, (C) TERT promoter alteration status, (D) tumour size (T stage), (E) CINSARC group and (F) chromosome 9p21.3 deletion. Corresponding P values are shown in each panel. Panels A-D include primary samples from the discovery and expansion cohorts; panels E and F are restricted to primary samples from the discovery cohort. G: Forest plot summarizing hazard ratios and 95% confidence intervals from a multivariable Cox proportional hazards model including most robust features significant in univariate survival analyses which were selected by bootstrap stability selection. Squares represent hazard ratio estimates and horizontal bars indicate 95% confidence intervals. Reference categories, sample sizes, hazard ratios, confidence intervals, and corresponding P values are shown. Model performance statistics are provided below the plot.

All candidate prognostic features were tested for multicollinearity and pairwise dependencies (Supplementary Table 6); none were collinear. Baseline metastatic status showed a significant moderate association with pTERT status (Supplementary Table 6). Given the cohort size, bootstrap-based variable-selection stability was assessed using LASSO-penalised Cox proportional hazards regression with 1,000 bootstrap resamples to identify the most robust candidate predictors for multivariable analysis. CINSARC class (96.3%), chr17q gain (79.3%), and baseline metastatic status (75.9%) emerged as the most stable predictors, each exceeding the pre-specified 70% selection threshold (Supplementary Table 7). Although pTERT was significant on univariate analysis, it did not meet the bootstrap stability threshold and was not carried forward to multivariable modelling. The proportional hazards assumption for the multivariable Cox model incorporating the three retained variables was confirmed using scaled Schoenfeld residuals (Supplementary Figure 6D). All three predictors: CINSARC class, chr17q gain, and baseline metastatic status retained independent prognostic significance for overall survival in the multivariable model (Figure 6G), together defining a genomic and clinical risk framework for CCS that has not previously been available for this diagnosis. To ensure our findings were not driven by inclusion of GI-CCS cases, we repeated univariate survival analysis after removing those cases from the cohort (*n*=4). All other factors remained significantly associated with OS (Supplementary Figure 6E-I) whereas pTERT was no longer statistically significant (p = 0.12), although Kaplan-Meier curves showed a similar trend (Supplementary Figure 6J), likely reflecting reduced statistical power.

To integrate these findings, we propose an evolutionary model of CCS in which a shared truncal fusion associates with a melanocytic lineage identity while divergent post-fusion copy-number trajectories determine clinical behaviour (Figure 7). Two distinct aneuploidy phenotypes emerge: a “loss-biased” cluster characterised by deletion of *CDKN2A* and *TP53* loss of heterozygosity, in which FGA correlates with progressive p53 pathway disruption; and a “gain-biased” cluster enriched for chr17q and chr1q copy-number gains, which associates with a transcriptional convergence on proliferative and glycolytic programmes. Superimposed chromosomal “catastrophe” events, including chromothripsis is seen in 50% of cases where genome sequencing data was available. Chromoplexy-mediated fusion formation, and whole-genome doubling correlate with subclonal diversification and, in CCS15, the case with longitudinal profiling, with metastatic progression.

**Figure 7:**
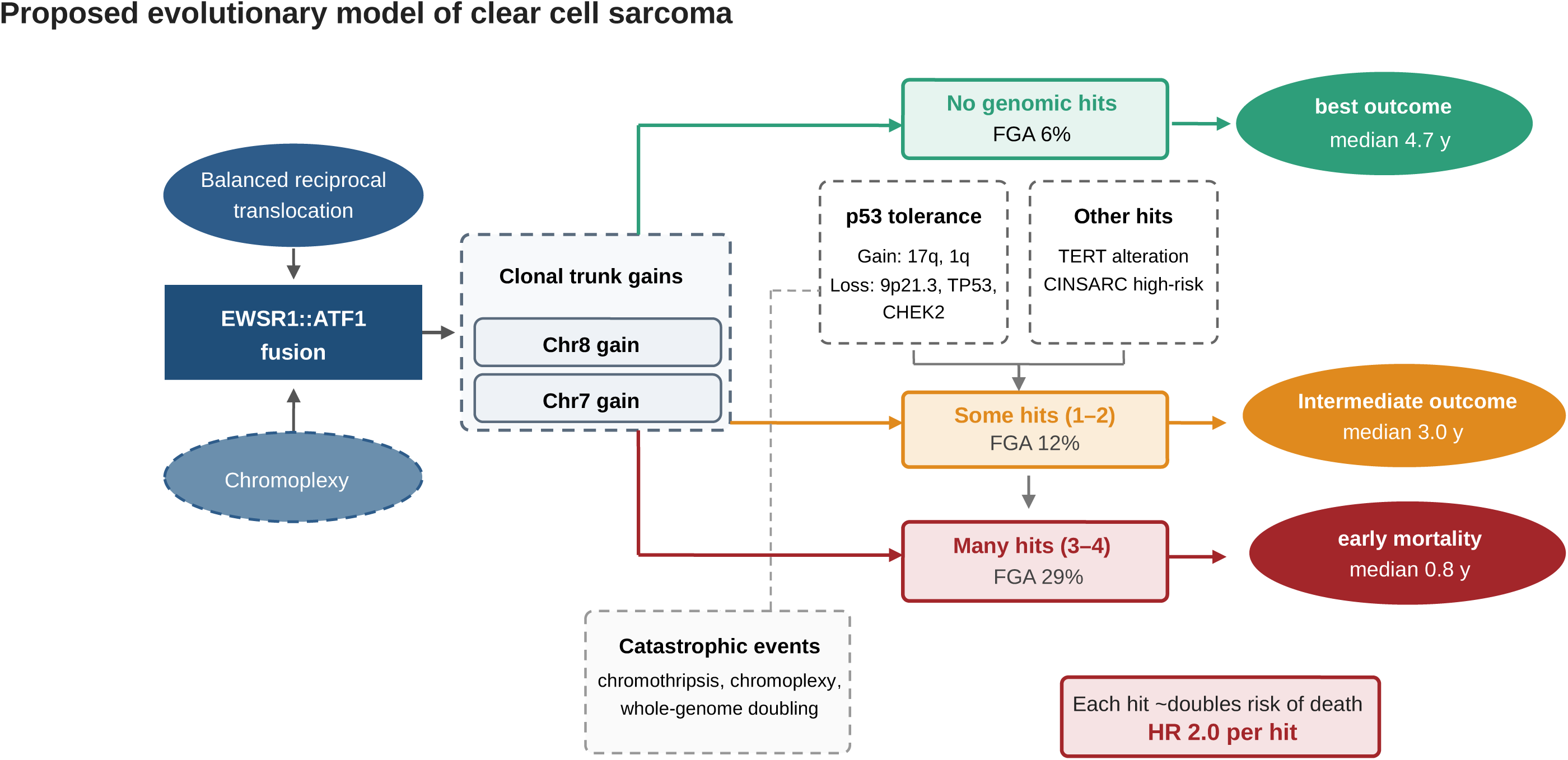
Evolutionary model of CCS in 37 primary tumours. All tumours carry the EWSR1::ATF1 fusion with clonal gains of chromosomes 8 and 7. Tumours then diverge by how many of four adverse alterations they acquire, with aneuploidy burden rising and survival falling across the three states. Counts are tumours per state; FGA, fraction of genome altered; survival by Firth-penalised Cox regression, age-adjusted.

## Discussion

CCS of soft tissue has long been considered a genomically uncomplicated fusion-driven sarcoma; the canonical driver *EWSR1::ATF1* arises as a truncal, clonal event in virtually all cases. Despite the involvement of alternative partner genes (*ATF1, CREB1, CREM*), all members of the CREB subfamily of bZIP domain transcription factors and appear functionally interchangeable in engaging the MITF/SOX10 melanocytic transcriptional programme (Möller et al., 2022). The fusion defines the disease, yet it does not explain why patients who carry it follow such varied clinical courses (Ikuta et al., 2023).

By integrating whole-genome, transcriptomic, evolutionary, immune and carefully curated clinical data across a multi-institutional cohort, we propose an evolutionary model in which the fusion fixes lineage and identity, while clinical behaviour is associated with subsequent genomic alterations (Figure 7). This has two practical consequences. CCS, in which histological grade carries no prognostic value (WHO Classification of Tumours Editorial Board, 2026) (Coindre, 2006) (Deyrup & Weiss, 2006), can be stratified by a small number of features measurable at diagnosis, and the same features point to therapeutic vulnerabilities that can now be tested.

The central feature of this model is where CCS sits on the aneuploidy spectrum. On a background of low point-mutation burden and near-diploid ploidy, CCS carries more aneuploidy than other fusion-driven sarcomas but far less than complex-karyotype sarcomas, and its altered genome is dominated by arm- and whole-chromosome-scale segments rather than the fragmented profiles of complex genome sarcomas. Clonal and subclonal change are of similar extent in CCS, whereas clonal change dominates in complex-karyotype sarcomas, indicating that CCS genomes keep diversifying after transformation, most plausibly through chromosome missegregation. This intermediate position fits models in which tumour fitness peaks within a narrow range of chromosomal instability, where moderate missegregation maximises clonal diversity while higher rates are selected against (Silk et al., 2013) (Laughney et al., 2015), and in which extreme instability is associated with improved outcome (Birkbak et al., 2011).

Sustained aneuploidy would normally engage p53-dependent checkpoints, and our data suggest that CCS reaches tolerance by two genomic routes to the same endpoint. In a smaller, “loss-biased” group, the TP53 axis is disrupted directly: 17p loss of heterozygosity associates with aneuploidy burden and co-occurs with loss of *CHEK2* and of 9p21.3, the CDKN2A/ARF locus. In the larger, “gain-biased” group, *TP53* wild-type tumours likely attenuate p53 through gene dosage, with 17q gains carrying *PPM1D* and 1q gains carrying *MDM4*, both known inhibitors of P53 activity (Milosevic et al., 2021) (Woodfield et al., 2021). The usual accelerants of extreme instability are uncommon: TP53 was altered in 5 of 37 primary tumours and whole-genome doubling occurred in 2 of 37 primary tumours. We propose that this dosage-level attenuation allows aneuploidy to be tolerated without the escalation that follows a complete loss of p53 function. Together with low immune infiltration, this may leave CCS in a therapeutic no-man’s land, buffered against DNA damage without being fragile, and poorly visible to T cells. This hypothesis raises the possibility that adding further incremental DNA damage would bring limited benefit, whereas the p53-intact majority may respond to strategies that remove the tolerance, such as MDM2/MDMX or PPM1D inhibition.

The order of events provides “genomic signposts” where intervention might act. Gains of chromosomes 8 and 7 are clonal trunk events in most tumours that carry them, whereas 17q gain is late in mutation time and frequently subclonal. Unlike in neuroblastoma, where 17q gain is an early event, in CCS it appears to mark progression and the emergence of aggressive subclones rather than initiation, and independent gains of chromosome 7 across longitudinal samples (Supplementary Figure 2G and H) indicate ongoing selection. Gain of 17q also raises the dosage of BIRC5 (survivin), which is overexpressed in tumours with this gain and can be targeted directly or through HDAC inhibition, as shown in Ewing sarcoma (Koppenhafer et al., 2025). Superimposed on this background, a subset of tumours undergo genome catastrophes, including chromoplexy-mediated fusion formation and chromothripsis; to our knowledge these have not been described in CCS, although they are known in other fusion-driven cancers (Anderson et al., 2018) (Baca et al., 2013). The enrichment of chromothriptic tumours for SBS13 suggests replication stress and error-prone repair (Maciejowski et al., 2020) (S. Liu et al., 2018), making this subset a candidate for replication-stress targeting. Because late subclonal events such as 17q gain may be missed in a single biopsy, longitudinal sampling of primary and metastatic disease will be needed to establish when high-risk subclones arise and whether they can be detected earlier, including in circulating tumour DNA.

Several of these alterations converge on *MYC*. *MGA* disruption, not previously described in CCS, together with chromosome 8 gain and enriched *MYC* transcriptional signatures, points to an underappreciated route to *MYC* pathway activation, with rRNA processing and ribosome biogenesis as the dominant downstream programmes. *MYC*-high CCS may therefore be susceptible to inhibition of Pol I transcription or early rRNA processing (Drygin et al., 2011) (Khot et al., 2019), a dependency that could also explain the limited activity of upstream inhibitors such as crizotinib (Schöffski et al., 2017). TERT promoter alterations, associated with adverse outcome here as in other sarcomas (Cross et al., 2022) (Demicco et al., 2018) (Y. Zhang et al., 2021), implicate telomere maintenance in progression (Gaspar et al., 2018). Despite heterogeneous copy-number and structural change, tumours converge on a limited number of transcriptional states that upregulate metabolic and proliferation pathways, which should simplify the design of biomarker-led studies.

These findings matter most for risk assessment. Alongside metastatic disease at diagnosis and large tumour size, established prognostic factors in CCS (H. Y. Wang et al., 2025) the CINSARC high-risk class and 17q gain were the most stable independent predictors of survival, and 17q gain remained associated with outcome independently of metastatic status and CINSARC class. Both features can be measured with clinically available methods, 17q gain by fluorescence in-situ hybridisation or sequencing and CINSARC by RNA-based testing. Before they guide practice, they need confirmation in independent, prospectively collected cohorts, ideally with uniform treatment data. If confirmed, they could be used to stratify entry into clinical trials, to intensify surveillance or adjuvant therapy for patients at high risk, and to spare patients at low risk from treatment of limited benefit.

The immune findings shape which therapies are potentially realistic. Most tumours show low immune infiltration with intact antigen presentation machinery, suggesting that the barrier lies in T-cell access and priming rather than antigen display. This favours approaches that supply specificity, such as T-cell receptor-based therapies, T-cell engagers or antibody–drug conjugates, over checkpoint blockade alone. In the separate companion spatial proteomics study, a proliferative tumour-cell state and an E2F-target module were associated with survival, mirroring the link between high aneuploidy and G2M and E2F programmes in our transcriptomes. Genome and proteome therefore point to a single proliferative axis, and testing whether it can be read from routine histology would bring risk assessment closer to the clinic.

Our observations in a unique multi-institutional cohort support an evolutionary model in which the *EWSR1* fusion initiates CCS, while acquired secondary alterations, accumulating on a background of tolerated aneuploidy, are associated with an increasingly unfavourable clinical course. The key genomic and transcriptomic features of primary tumours are recapitulated in CCS cell lines, which now allow the central predictions to be tested: that dosage-attenuated p53 can be reactivated, that MYC-high tumours depend on ribosome biogenesis, and that pushing chromosome missegregation beyond tolerance is selectively lethal. Combined with prospective sampling of primary and metastatic disease, these models provide a foundation for drug discovery in this ultra-rare cancer.

### Limitations of the study and future work

Clinical and follow-up data were incomplete for the expansion cohort, so survival analyses were restricted to patients with available data. The low indel burden of CCS limited the power to assign indel signatures, which should therefore be regarded as exploratory. Copy-number profiles capture the outcome of chromosomal instability (CIN) rather than its rate, so our inferences about ongoing instability and its restraint are indirect. The subclonal fraction of genome altered was estimated from single samples, which underestimates subclonality in all tumours, probably most in complex, genome-doubled tumours such as UPS; this could exaggerate the contrast between CCS and complex-karyotype sarcomas. Differences in sequencing depth between our cohort and the Genomics England comparator data may also affect subclonal calling. Even so, a substantial subclonal burden together with a distinct, large-segment pattern of alteration is consistent with ongoing CIN in CCS. Prognostic features were selected and modelled in the same cohort, so survival estimates are exploratory, and the CINSARC high-risk class and 17q gain require validation in an independent series. Future work should measure chromosome missegregation directly in CCS cell lines and tumour sections and test the tolerance and therapeutic hypotheses functionally. Larger cohorts with multi-region, longitudinal and single-cell data will be needed to resolve subclonal trajectories and determine how they shape progression and treatment resistance.

## Acknowledgements

We would like to thank the patients and their friends & families from across the world for the generous donation of their time, feedback and materials which enabled this study.

This study was funded by Edward Showler Foundation (https://www.edwardshowlerfoundation.com), Transcend Sarcoma (https://www.transcendsarcoma.com) and Sarcoma UK (https://sarcoma.org.uk). N.P. has received funding from Cancer Research UK (Award number 18387) and holds a Cancer Research UK Career Establishment award (RCCCEA-Nov23/100003). T.B. received Jean Sharks Pathological Society Clinical PhD fellowship grant. C.D holds a Sarcoma UK-funded studentship (Sayako Grace Robinson studentship - SUK09.2022). Provision of patients’ samples was made possible through the Royal National Orthopaedic Hospital Pathology Department, and The Bone Cancer Research Trust.

D. K. received support from the Else Kröner-Fresenius-Foundation through the University Medicine Essen Medical Scientist Academy program. The project was further supported by the Stiftung Sarkomforschung (Sarcoma Research Foundation) and the www.sarkomtour.de fundraising initiative.

S.W., T.L., and P.V.L. were supported by the Francis Crick Institute, which receives its core funding from Cancer Research UK (CC2008), the UK Medical Research Council (CC2008) and the Wellcome Trust (CC2008). P.V.L. is a CPRIT Scholar in Cancer Research and acknowledges CPRIT grant support (RR210006).

This work was also supported through the HEROES-AYA (Heterogeneity, Evolution, and Resistance in Oncogenic Fusion Gene-Expressing Sarcomas Affecting Adolescents and Young Adults) consortium within the National Decade Against Cancer of the German Federal Ministry of Education and Research (01KD2207).

We are grateful to the UCL/UCLH biobank for health and disease (RNOH satellite) and Queen square brain bank for Neurological Disorders at UCL. We are also grateful to the healthcare workers who cared for the patients. Research infrastructure at Francis Crick Institute was supported by the Wellcome Trust, Cancer Research UK, and Medical Research Council. We thank the Translational Technology Platforms at the UCL Cancer Institute for their technical support and assistance in this work. These facilities are supported by the Cancer Research UK City of London Centre Award [CTRQQR-2021\100004]. We acknowledge the support of UCL Genomics (RRID:SCR_027010) for methylation profiling of our samples.

We want to thank Dr. Maxime Tarabichi (Group Leader, Interuniversity Institute of Bioinformatics in Brussels, Brussels, Belgium) and Dr. Jonas Demeulemeester (Group Leader, VIB – KU Leuven Center for Cancer Biology, Leuven, Belgium) and his team for their technical expertise and support in the analysis. We also want to thank Dr. Jianan Chen (UCL Cancer Institute) for his valuable feedback and expertise on ploidy discussion.

We gratefully acknowledge the participants of the National Genomic Research Library (NGRL), whose contributions made this research possible. Secure access to the NGRL under project ID RR1215 was provided by Genomics England, which delivers the NGRL in partnership with NHS England, and is wholly owned by the UK Department of Health and Social Care. The NGRL contains participants’ health data collected by the NHS as part of their care, along with samples and data from their participation in research, for which fully informed consent has been obtained. This includes genomic and clinical data provided through the NHS Genomic Medicine Service, as well as data obtained through research studies, including the 100,000 Genomes Project and the Generation Study, both of which are delivered in partnership with the NHS, and from other research cohorts involving external collaborators.

For the provision of cell lines, we would like to thank the HUSARC cooperative & Dr Susanne Grunewalk (West German Cancer Center, Germany) for the CCS-SN1 line, Dr Beate Rinner & Katharina Meditz (Medical University of Graz, Austria) for the MUG-Lucifer lines, Dr Hidetatsu Otani (Osaka University, Japan) for the Hewga-CCS lines, ATCC (American Type Culture Collection) for the SUCCS-1 line, Dr Christian Britschgi (University Hospital Zurich, Switzerland) for the USZ-CCS-1 line, Dr Jonathan Fletcher (Brigham and Women’s Hospital, USA) for the CCS24 & CCS210 lines.

During the preparation of this manuscript, the authors used Claude (Opus 5.5; Anthropic) to improve the language and readability of the Discussion section, to check analysis code written by the authors, and to assist in arranging author-generated panels into composite figures and to adjust font size and style in author-created figures for consistency. The tool did not generate or modify any data or image content. The authors reviewed and edited all content and take full responsibility for the publication.

## Data availability

Genomic data generated in this study will be deposited in the European Genome-phenome Archive (EGA) as well as in German Human Genome-Phenome Archive (GHGA) and will be made available through controlled access upon publication. The accession number will be provided upon acceptance of the manuscript. Data from the National Genomic Research Library (NGRL) (The National Genomic Research Library, 2024) used in this research are available within the secure Genomics England Research Environment. Access to NGRL data is restricted to adhere to consent requirements and protect participant privacy. Data used in this research include cancer sarcoma data from the Genomics England 100,000 Genomes Project, version 16, released October 13, 2022. Data used in this research include the following metrics derived from whole-genome sequencing of sarcomas in the 100,000 Genomes Project: fraction of genome altered (FGA), subclonal FGA, number of subclonal segments, and the cumulative distributions of clonal and subclonal segments for each sarcoma subtype analysed. Access to NGRL data is provided to approved researchers who are members of the Genomics England Research Network, subject to institutional access agreements and research project approval under participant-led governance. For more information on data access, visit: https://www.genomicsengland.co.uk/research

## Code Availability

Code used for analysis and generation of figures will be made publicly available upon acceptance.

## References

Anderson, N. D., De Borja, R., Young, M. D., Fuligni, F., Rosic, A., Roberts, N. D., Hajjar, S., Layeghifard, M., Novokmet, A., Kowalski, P. E., Anaka, M., Davidson, S., Zarrei, M., Said, B. I., Christine Schreiner, L., Marchand, R., Sitter, J., Gokgoz, N., Brunga, L., … Shlien, A. (2018). Rearrangement bursts generate canonical gene fusions in bone and soft tissue tumors. Science, 361(6405). 10.1126/science.aam8419

Antonescu, C. R., Tschernyavsky, S. J., Woodruff, J. M., Jungbluth, A. A., Brennan, M. F., & Ladanyi, M. (2002). Molecular Diagnosis of Clear Cell Sarcoma Detection of EWS-ATF1 and MITF-M Transcripts and Histopathological and Ultrastructural Analysis of 12 Cases.

Baca, S. C., Prandi, D., Lawrence, M. S., Mosquera, J. M., Romanel, A., Drier, Y., Park, K., Kitabayashi, N., MacDonald, T. Y., Ghandi, M., Van Allen, E., Kryukov, G. V., Sboner, A., Theurillat, J. P., Soong, T. D., Nickerson, E., Auclair, D., Tewari, A., Beltran, H., … Garraway, L. A. (2013). Punctuated evolution of prostate cancer genomes. Cell, 153(3), 666–677. 10.1016/j.cell.2013.03.021

Baker, T. M., Lai, S., Lynch, A. R., Lesluyes, T., Yan, H., Ogilvie, H. A., Verfaillie, A., Dentro, S., Bowes, A. L., Pillay, N., Flanagan, A. M., Swanton, C., Spellman, P. T., Tarabichi, M., & Van Loo, P. (2024). The History of Chromosomal Instability in Genome-Doubled Tumors. Cancer Discovery, 14(10), 1810–1822. 10.1158/2159-8290.CD-23-1249

Barbie, D. A., Tamayo, P., Boehm, J. S., Kim, S. Y., Moody, S. E., Dunn, I. F., Schinzel, A. C., Sandy, P., Meylan, E., Scholl, C., Fröhling, S., Chan, E. M., Sos, M. L., Michel, K., Mermel, C., Silver, S. J., Weir, B. A., Reiling, J. H., Sheng, Q., … Hahn, W. C. (2009). Systematic RNA interference reveals that oncogenic KRAS-driven cancers require TBK1. Nature, 462(7269), 108–112. 10.1038/nature08460

Barry, K. H., Mohanty, K., Erickson, P. A., Wang, D., Shi, J., Rose, G., Cellini, A., Clark, K., Ambulos, N., Yin, J., Yan, L., Poulin, M., Meyer, A., Zhang, Y., Bentzen, S. M., Burke, A., Hussain, A., & Berndt, S. I. (2021). Myc dna methylation in prostate tumor tissue is associated with gleason score. Genes, 12(1), 1–23. 10.3390/genes12010012

Bielski, C. M., Zehir, A., Penson, A. V., Donoghue, M. T. A., Chatila, W., Armenia, J., Chang, M. T., Schram, A. M., Jonsson, P., Bandlamudi, C., Razavi, P., Iyer, G., Robson, M. E., Stadler, Z. K., Schultz, N., Baselga, J., Solit, D. B., Hyman, D. M., Berger, M. F., & Taylor, B. S. (2018). Genome doubling shapes the evolution and prognosis of advanced cancers. Nature Genetics, 50(8), 1189–1195. 10.1038/s41588-018-0165-1

Birkbak, N. J., Eklund, A. C., Li, Q., McClelland, S. E., Endesfelder, D., Tan, P., Tan, I. B., Richardson, A. L., Szallasi, Z., & Swanton, C. (2011). Paradoxical relationship between chromosomal instability and survival outcome in cancer. Cancer Research, 71(10), 3447–3452. 10.1158/0008-5472.CAN-10-3667

Bridge, U. A., Sreekantaiah, C., Ames, ], Neff, R., & Sandberg, A. A. (1991). Cytogenetic Findings in Clear Cell Sarcoma of Tendons and Aponeuroses Malignant Melanoma of Soft Parts.

Brohl, A. S., Solomon, D. A., Chang, W., Wang, J., Song, Y., Sindiri, S., Patidar, R., Hurd, L., Chen, L., Shern, J. F., Liao, H., Wen, X., Gerard, J., Kim, J. S., Lopez Guerrero, J. A., Machado, I., Wai, D. H., Picci, P., Triche, T., … Khan, J. (2014). The Genomic Landscape of the Ewing Sarcoma Family of Tumors Reveals Recurrent STAG2 Mutation. PLoS Genetics, 10(7). 10.1371/journal.pgen.1004475

Brown, A. D., Lopez-Terrada, D., Denny, C., & Lee, K. A. (1995). Promoters containing ATF-binding sites are de-regulated in cells that express the EWS/ATF1 oncogene. Oncogene, 10(9), 1749–1756.

Butters, T., Oukrif, D., Tannirandorn, P., van Belzen, I. A., Chen, J., Davies, C., Lin, R., De Noon, S., Taher, D., Cutler, C., Markov, M., Tollit, A., Oumlil, F., Steele, C. D., Fittall, M., Guo, Y., Cortés-Ciriano, I., Van Loo, P., Flanagan, A. M., … Tarabichi, M. (2026). Improving computational tumor ploidy estimation in complex cancer genomes through flow-cytometry-guided calibration. Cell Reports Methods, 6(10). 10.1016/j.crmeth.2026.101570

Canisius, S., Martens, J. W. M., & Wessels, L. F. A. (2016). A novel independence test for somatic alterations in cancer shows that biology drives mutual exclusivity but chance explains most co-occurrence. Genome Biology, 17(1). 10.1186/s13059-016-1114-x

Carvalho, C. M. B., & Lupski, J. R. (2016). Mechanisms underlying structural variant formation in genomic disorders. In Nature Reviews Genetics (Vol. 17, Number 4, pp. 224–238). Nature Publishing Group. 10.1038/nrg.2015.25

Chalmers, Z. R., Connelly, C. F., Fabrizio, D., Gay, L., Ali, S. M., Ennis, R., Schrock, A., Campbell, B., Shlien, A., Chmielecki, J., Huang, F., He, Y., Sun, J., Tabori, U., Kennedy, M., Lieber, D. S., Roels, S., White, J., Otto, G. A., … Frampton, G. M. (2017). Analysis of 100,000 human cancer genomes reveals the landscape of tumor mutational burden. Genome Medicine, 9(1). 10.1186/s13073-017-0424-2

Chibon, F., Lagarde, P., Salas, S., Pérot, G., Brouste, V., Tirode, F., Lucchesi, C., De Reynies, A., Kauffmann, A., Bui, B., Terrier, P., Bonvalot, S., Le Cesne, A., Vince-Ranchère, D., Blay, J. Y., Collin, F., Guillou, L., Leroux, A., Coindre, J. M., & Aurias, A. (2010). Validated prediction of clinical outcome in sarcomas and multiple types of cancer on the basis of a gene expression signature related to genome complexity. Nature Medicine, 16(7), 781–787. 10.1038/nm.2174

Chowdhury, M. A. R., Haq, M. M., Lee, J. H., & Jeong, S. (2024). Multi-faceted regulation of CREB family transcription factors. In Frontiers in Molecular Neuroscience (Vol. 17). Frontiers Media SA. 10.3389/fnmol.2024.1408949

Chung, E. B., & Enzinger, F. M. (1983). Malignant melanoma of soft parts A reassessment of clear cell sarcoma*. In The American Journal of Surgical Pathology (Vol. 7).

Coindre, J.-M. (2006). Grading of Soft Tissue Sarcomas: Review and Update. Archives of Pathology & Laboratory Medicine, 130(10), 1448–1453. 10.5858/2006-130-1448-GOSTSR

Cornish, A. J., Gruber, A. J., Kinnersley, B., Chubb, D., Frangou, A., Caravagna, G., Noyvert, B., Lakatos, E., Wood, H. M., Thorn, S., Culliford, R., Arnedo-Pac, C., Househam, J., Cross, W., Sud, A., Law, P., Leathlobhair, M. N., Hawari, A., Woolley, C., … Houlston, R. S. (2024). The genomic landscape of 2,023 colorectal cancers. Nature, 633(8028), 127–136. 10.1038/s41586-024-07747-9

Cortés-Ciriano, I., Lee, J. J.-K., Xi, R., Jain, D., Jung, Y. L., Yang, L., Gordenin, D., Klimczak, L. J., Zhang, C.-Z., Pellman, D. S., Akdemir, K. C., Alvarez, E. G., Baez-Ortega, A., Beroukhim, R., Boutros, P. C., Bowtell, D. D. L., Brors, B., Burns, K. H., Campbell, P. J., … von Mering, C. (2020). Comprehensive analysis of chromothripsis in 2,658 human cancers using whole-genome sequencing. Nature Genetics, 52(3), 331–341. 10.1038/s41588-019-0576-7

Cortes-Ciriano, I., Steele, C. D., Piculell, K., Al-Ibraheemi, A., Eulo, V., Bui, M. M., Chatzipli, A., Dickson, B. C., Borcherding, D. C., Feber, A., Galor, A., Hart, J., Jones, K. B., Jordan, J. T., Kim, R. H., Lindsay, D., Miller, C., Nishida, Y., Proszek, P. Z., … Miller, D. T. (2023). Genomic Patterns of Malignant Peripheral Nerve Sheath Tumor (MPNST) Evolution Correlate with Clinical Outcome and Are Detectable in Cell-Free DNA. Cancer Discovery, 13(3), 654–671. 10.1158/2159-8290.CD-22-0786

Cross, W., Lyskjær, I., Lesluyes, T., Hargreaves, S., Strobl, A. C., Davies, C., Waise, S., Hames-Fathi, S., Oukrif, D., Ye, H., Amary, F., Tirabosco, R., Gerrand, C., Baker, T., Barnes, D., Steele, C., Alexandrov, L., Bond, G., Cool, P., … Flanagan, A. M. (2022). A genetic model for central chondrosarcoma evolution correlates with patient outcome. Genome Medicine, 14(1). 10.1186/s13073-022-01084-0

Davis, I. J., Kim, J. J., Ozsolak, F., Widlund, H. R., Rozenblatt-Rosen, O., Granter, S. R., Du, J., Fletcher, J. A., Denny, C. T., Lessnick, S. L., Linehan, W. M., Kung, A. L., & Fisher, D. E. (2006). Oncogenic MITF dysregulation in clear cell sarcoma: Defining the MiT family of human cancers. Cancer Cell, 9(6), 473–484. 10.1016/j.ccr.2006.04.021

Demicco, E. G., Wani, K., Ingram, D., Wagner, M., Maki, R. G., Rizzo, A., Meeker, A., Lazar, A. J., & Wang, W. L. (2018). TERT promoter mutations in solitary fibrous tumour. Histopathology, 73(5), 843–851. 10.1111/his.13703

Deyrup, A. T., & Weiss, S. W. (2006). Grading of soft tissue sarcomas: the challenge of providing precise information in an imprecise world. Histopathology, 48(1), 42–50. 10.1111/j.1365-2559.2005.02288.x

Drygin, D., Lin, A., Bliesath, J., Ho, C. B., O’Brien, S. E., Proffitt, C., Omori, M., Haddach, M., Schwaebe, M. K., Siddiqui-Jain, A., Streiner, N., Quin, J. E., Sanij, E., Bywater, M. J., Hannan, R. D., Ryckman, D., Anderes, K., & Rice, W. G. (2011). Targeting RNA polymerase I with an oral small molecule CX-5461 inhibits ribosomal RNA synthesis and solid tumor growth. Cancer Research, 71(4), 1418–1430. 10.1158/0008-5472.CAN-10-1728

Enzinger, F. M. (1965). Clear-cell sarcoma of tendons and aponeuroses. An analysis of 21 cases. Cancer, 18(9), 1163–1174. 10.1002/1097-0142(196509)18:9<1163::AID-CNCR2820180916>3.0.CO;2-0

Epstein, A. L., Martin, A. O., & Kempson, R. (1983). Use of a newly established human cell line (SU-CCS-1) to demonstrate the relationship of clear cell sarcoma to malignant melanoma. Cancer Research.

Gaspar, T. B., Sá, A., Lopes, J. M., Sobrinho-Simões, M., Soares, P., & Vinagre, J. (2018). Telomere maintenance mechanisms in cancer. In Genes (Vol. 9, Number 5). MDPI AG. 10.3390/genes9050241

Graadt Van Roggen, J. F., Mooi, W. J., & Hogendoorn, P. C. W. (1998). Clear cell sarcoma of tendons and aponeuroses (malignant melanoma of soft parts) and cutaneous melanoma: Exploring the histogenetic relationship between these two clinicopathological entities. In Journal of Pathology (Vol. 186, Number 1, pp. 3–7). John Wiley and Sons Ltd. 10.1002/(SICI)1096-9896(199809)186:1<3::AID-PATH153>3.0.CO;2-V

Hänzelmann, S., Castelo, R., & Guinney, J. (2013). GSVA: gene set variation analysis for microarray and RNA-Seq data. In BMC Bioinformatics (Vol. 14). http://www.biomedcentral.com/1471-2105/14/7http://www.bioconductor.org.Background

Hiraga, H., Nojima, T., Abe, S., Yamashiro, K., Yamawaki, S., Kaneda, K., & Nagashima, K. (1997). Establishment of a new continuous clear cell sarcoma cell line. Virchows Archiv, 431(1), 45–51. 10.1007/s004280050068

Horak, P., Heining, C., Kreutzfeldt, S., Hutter, B., Mock, A., Hüllein, J., Fröhlich, M., Uhrig, S., Jahn, A., Rump, A., Gieldon, L., Möhrmann, L., Hanf, D., Teleanu, V., Heilig, C. E., Lipka, D. B., Allgäuer, M., Ruhnke, L., Laßmann, A., … Fröhling, S. (2021). Comprehensive Genomic and Transcriptomic Analysis for Guiding Therapeutic Decisions in Patients with Rare Cancers. Cancer Discovery, 11(11), 2780–2795. 10.1158/2159-8290.CD-21-0126

Hugo, W., Zaretsky, J. M., Sun, L., Song, C., Moreno, B. H., Hu-Lieskovan, S., Berent-Maoz, B., Pang, J., Chmielowski, B., Cherry, G., Seja, E., Lomeli, S., Kong, X., Kelley, M. C., Sosman, J. A., Johnson, D. B., Ribas, A., & Lo, R. S. (2016). Genomic and Transcriptomic Features of Response to Anti-PD-1 Therapy in Metastatic Melanoma. Cell, 165(1), 35–44. 10.1016/j.cell.2016.02.065

Hurlin, P. J., Eirìkur, E., Steingrìmsson, S., Copeland, N. G., Jenkins, N. A., & Eisenman, R. N. (1999). Mga, a dual-specificity transcription factor that interacts with Max and contains a T-domain DNA-binding motif. In The EMBO Journal (Vol. 18, Number 24).

Ibrahim, R. M., Steenstrup Jensen, S., & Juel, J. (2018). Clear cell sarcoma-A review. In Journal of Orthopaedics (Vol. 15, Number 4, pp. 963–966). Reed Elsevier India Pvt. Ltd. 10.1016/j.jor.2018.08.039

Ikuta, K., Nishida, Y., Imagama, S., Tanaka, K., & Ozaki, T. (2023). The current management of clear cell sarcoma. In Japanese Journal of Clinical Oncology (Vol. 53, Number 10, pp. 899–904). Oxford University Press. 10.1093/jjco/hyad083

Johnson, W. E., Li, C., & Rabinovic, A. (2007). Adjusting batch effects in microarray expression data using empirical Bayes methods. Biostatistics, 8(1), 118–127. 10.1093/biostatistics/kxj037

Jones, R. L., Constantinidou, A., Thway, K., Ashley, S., Scurr, M., Al-Muderis, O., Fisher, C., Antonescu, C. R., D’Adamo, D. R., Keohan, M. L., Maki, R. G., & Judson, I. R. (2011). Chemotherapy in clear cell sarcoma. Medical Oncology, 28(3), 859–863. 10.1007/s12032-010-9502-7

Karner, C., Anders, I., Vejzovic, D., Szkandera, J., Scheipl, S., Deutsch, A. J. A., Weiss, L., Vierlinger, K., Kolb, D., Kühberger, S., Heitzer, E., Habisch, H., Zhang, F., Madl, T., Reininger-Gutmann, B., Liegl-Atzwanger, B., & Rinner, B. (2023). Targeting epigenetic features in clear cell sarcomas based on patient-derived cell lines. Journal of Translational Medicine, 21(1). 10.1186/s12967-022-03843-4

Kaufmann, T. L., Petkovic, M., Watkins, T. B. K., Colliver, E. C., Laskina, S., Thapa, N., Minussi, D. C., Navin, N., Swanton, C., Van Loo, P., Haase, K., Tarabichi, M., & Schwarz, R. F. (2022). MEDICC2: whole-genome doubling aware copy-number phylogenies for cancer evolution. Genome Biology, 23(1). 10.1186/s13059-022-02794-9

Kawai, A., Hosono, A., Nakayama, R., Matsumine, A., Matsumoto, S., Ueda, T., Tsuchiya, H., Beppu, Y., Morioka, H., & Yabe, H. (2007). Clear cell sarcoma of tendons and aponeuroses: A study of 75 patients. Cancer, 109(1), 109–116. 10.1002/cncr.22380

Keung, E. Z., Burgess, M., Salazar, R., Parra, E. R., Rodrigues-Canales, J., Bolejack, V., Van Tine, B. A., Schuetze, S. M., Attia, S., Riedel, R. F., Hu, J., Okuno, S. H., Priebat, D. A., Movva, S., Davis, L. E., Reed, D. R., Reuben, A., Roland, C. L., Reinke, D., … Tawbi, H. A. (2020). Correlative analyses of the SARC028 trial reveal an association between sarcoma-associated immune infiltrate and response to pembrolizumab. Clinical Cancer Research, 26(6), 1258–1266. 10.1158/1078-0432.CCR-19-1824

Khan, A. A., Aptekmann, A. A., Min, D., Povelaitis, M. C., Lee, S., & Ryu, B. (2025). The CoREST Complex Regulates Alternative Splicing by the Transcriptional Regulation of RNA Processing Genes in Melanoma Cells. Cells, 14(21). 10.3390/cells14211699

Khot, A., Brajanovski, N., Cameron, D. P., Hein, N., Maclachlan, K. H., Sanij, E., Lim, J., Soong, J., Link, E., Blombery, P., Thompson, E. R., Fellowes, A., Sheppard, K. E., McArthur, G. A., Pearson, R. B., Hannan, R. D., Poortinga, G., & Harrison, S. J. (2019). First-in-human RNA polymerase I transcription inhibitor CX-5461 in patients with advanced hematologic cancers: Results of a phase I dose-escalation study. Cancer Discovery, 9(8), 1036–1049. 10.1158/2159-8290.CD-18-1455

Koppenhafer, S. L., Geary, E. L., Thomas, M. V., Croushore, E. E., Zimmerman, J. A. O., Gedminas, J. M., Quelle, D. E., Dodd, R. D., & Gordon, D. J. (2025). Histone Deacetylase Inhibitors Target DNA Replication Regulators and Replication Stress in Ewing Sarcoma Cells. Cancer Research Communications, 5(6), 1034–1048. 10.1158/2767-9764.CRC-25-0058

Korotkevich, G., Sukhov, V., Budin, N., Shpak, B., Artyomov, M. N., & Sergushichev, A. (2016). Fast gene set enrichment analysis. 10.1101/060012

Lafita-Navarro, M. C., Kim, M., Borenstein-Auerbach, N., Venkateswaran, N., Hao, Y. H., Ray, R., Brabletz, T., Scaglioni, P. P., Shay, J. W., & Conacci-Sorrell, M. (2018). The aryl hydrocarbon receptor regulates nucleolar activity and protein synthesis in MYC-expressing cells. Genes and Development, 32(19–20), 1303–1308. 10.1101/GAD.313007.118

Laughney, A. M., Elizalde, S., Genovese, G., & Bakhoum, S. F. (2015). Dynamics of Tumor Heterogeneity Derived from Clonal Karyotypic Evolution. Cell Reports, 12(5), 809–820. 10.1016/j.celrep.2015.06.065

Lee, C. J., Modave, E., Boeckx, B., Stacchiotti, S., Rutkowski, P., Blay, J. Y., Debiec-Rychter, M., Sciot, R., Lambrechts, D., Wozniak, A., & Schöffski, P. (2021). Histopathological and molecular profiling of clear cell sarcoma and correlation with response to crizotinib: An exploratory study related to EORTC 90101 “CREATE” trial. Cancers, 13(23). 10.3390/cancers13236057

Li, A. B., Jiang, B. J., Wang, H. H., Yang, Y. S., Zhang, X. B., Lan, G. H., & Shu, W. Bin. (2019). Prognostic factors for survival in patients with clear cell sarcoma: An analysis of the Surveillance, Epidemiology, and End Results (SEER) database. Medical Science Monitor, 25, 6939–6945. 10.12659/MSM.916705

Li, J., Chen, C., Liu, W., Liu, S., Hu, W., Gao, X., Liu, G., Li, D., Ding, Y., Wen, X., Zhang, X., Hou, Y., Zhang, X., Li, B., Zhang, X., & Zhang, X. (2021). Whole-exome sequencing in clear cell sarcoma of soft tissue uncovers novel prognostic categorization and drug targets. Clinical and Translational Medicine, 11(12). 10.1002/ctm2.640

Liberzon, A., Birger, C., Thorvaldsdóttir, H., Ghandi, M., Mesirov, J. P., & Tamayo, P. (2015). The Molecular Signatures Database Hallmark Gene Set Collection. Cell Systems, 1(6), 417–425. 10.1016/j.cels.2015.12.004

Liu, H., Zhang, H., Zhang, C., Liao, Z., Li, T., Yang, T., Zhang, G., & Yang, J. (2022). Pan-Soft Tissue Sarcoma Analysis of the Incidence, Survival, and Metastasis: A Population-Based Study Focusing on Distant Metastasis and Lymph Node Metastasis. Frontiers in Oncology, 12. 10.3389/fonc.2022.890040

Liu, S., Kwon, M., Mannino, M., Yang, N., Renda, F., Khodjakov, A., & Pellman, D. (2018). Nuclear envelope assembly defects link mitotic errors to chromothripsis. Nature, 561(7724), 551–555. 10.1038/s41586-018-0534-z

Maciejowski, J., Chatzipli, A., Dananberg, A., Chu, K., Toufektchan, E., Klimczak, L. J., Gordenin, D. A., Campbell, P. J., & de Lange, T. (2020). APOBEC3-dependent kataegis and TREX1-driven chromothripsis during telomere crisis. Nature Genetics, 52(9), 884–890. 10.1038/s41588-020-0667-5

Mermel, C. H., Schumacher, S. E., Hill, B., Meyerson, M. L., Beroukhim, R., & Getz, G. (2011). GISTIC2.0 facilitates sensitive and confident localization of the targets of focal somatic copy-number alteration in human cancers. Genome Biology, 12(4). 10.1186/gb-2011-12-4-r41

Milosevic, J., Treis, D., Fransson, S., Gallo-oller, G., Sveinbjörnsson, B., Eissler, N., Tanino, K., Sakaguchi, K., Martinsson, T., Wickström, M., Kogner, P., & Johnsen, J. I. (2021). Ppm1d is a therapeutic target in childhood neural tumors. Cancers, 13(23). 10.3390/cancers13236042

Mitelman F, J. B. M. F. and P. K. (Eds. ). (2026). Mitelman Database of Chromosome Aberrations and Gene Fusions in Cancer.

Möller, E., Praz, V., Rajendran, S., Dong, R., Cauderay, A., Xing, Y. H., Lee, L., Fusco, C., Broye, L. C., Cironi, L., Iyer, S., Rengarajan, S., Awad, M. E., Naigles, B., Letovanec, I., Ormas, N., Finzi, G., La Rosa, S., Sessa, F., … Riggi, N. (2022). EWSR1-ATF1 dependent 3D connectivity regulates oncogenic and differentiation programs in Clear Cell Sarcoma. Nature Communications, 13(1). 10.1038/s41467-022-29910-4

Morales, J., Pujar, S., Loveland, J. E., Astashyn, A., Bennett, R., Berry, A., Cox, E., Davidson, C., Ermolaeva, O., Farrell, C. M., Fatima, R., Gil, L., Goldfarb, T., Gonzalez, J. M., Haddad, D., Hardy, M., Hunt, T., Jackson, J., Joardar, V. S., … Murphy, T. D. (2022). A joint NCBI and EMBL-EBI transcript set for clinical genomics and research. Nature, 604(7905), 310–315. 10.1038/s41586-022-04558-8

Myers, M. A., Arnold, B. J., Bansal, V., Balaban, M., Mullen, K. M., Zaccaria, S., & Raphael, B. J. (2024). HATCHet2: clone- and haplotype-specific copy number inference from bulk tumor sequencing data. Genome Biology, 25(1). 10.1186/s13059-024-03267-x

Nik-Zainal, S., Van Loo, P., Wedge, D. C., Alexandrov, L. B., Greenman, C. D., Lau, K. W., Raine, K., Jones, D., Marshall, J., Ramakrishna, M., Shlien, A., Cooke, S. L., Hinton, J., Menzies, A., Stebbings, L. A., Leroy, C., Jia, M., Rance, R., Mudie, L. J.,… Campbell, P. J. (2012). The life history of 21 breast cancers. Cell, 149(5), 994–1007. 10.1016/j.cell.2012.04.023

P Dal Cin, H. V. den B. P. P. (1999). Epithelioid Sarcoma of the Proximal Type with Complex Karyotype Including i(8q). Cancer Genetics and Cytogenetics, 1(114), 80–82.

Panagopoulos, I., Mertens, F., Dêbiec-Rychter, M., Isaksson, M., Limon, J., Kardas, I., Domanski, H. A., Sciot, R., Perek, D., Crnalic, S., Larsson, O., & Mandahl, N. (2002). Molecular genetic characterization of the EWS/ATF1 fusion gene in clear cell sarcoma of tendons and aponeuroses. International Journal of Cancer, 99(4), 560–567. 10.1002/ijc.10404

Panza, E., Ozenberger, B. B., Straessler, K. M., Barrott, J. J., Li, L., Wang, Y., Xie, M., Boulet, A., Titen, S. W. A., Mason, C. C., Lazar, A. J., Ding, L., Capecchi, M. R., & Jones, K. B. (2021). The clear cell sarcoma functional genomic landscape. Journal of Clinical Investigation, 131(15). 10.1172/JCI146301

Peacock, T., Heather, J. M., Ronel, T., & Chain, B. (2021). Decombinator V4: An improved AIRR compliant-software package for T-cell receptor sequence annotation? Bioinformatics, 37(6), 876–878. 10.1093/bioinformatics/btaa758

Rasmussen, S. V., Wozniak, A., Lathara, M., Goldenberg, J. M., Samudio, B. M., Bickford, L. R., Nagamori, K., Wright, H., Woods, A. D., Chauhan, S., Lee, C. J., Rudzinski, E. R., Swift, M. K., Kondo, T., Fisher, D. E., Imyanitov, E., Machado, I., Llombart-Bosch, A., Andrulis, I. L., … Keller, C. (2023). Functional genomics of human clear cell sarcoma: genomic, transcriptomic and chemical biology landscape for clear cell sarcoma. British Journal of Cancer, 128(10), 1941–1954. 10.1038/s41416-023-02222-0

Ritchie, M. E., Phipson, B., Wu, D., Hu, Y., Law, C. W., Shi, W., & Smyth, G. K. (2015). Limma powers differential expression analyses for RNA-sequencing and microarray studies. Nucleic Acids Research, 43(7), e47. 10.1093/nar/gkv007

Schöffski, P., Wozniak, A., Stacchiotti, S., Rutkowski, P., Blay, J. Y., Lindner, L. H., Strauss, S. J., Anthoney, A., Duffaud, F., Richter, S., Grünwald, V., Leahy, M. G., Reichardt, P., Sufliarsky, J., van der Graaf, W. T., Sciot, R., Debiec-Rychter, M., Van Cann, T., Marréaud, S., … Bauer, S. (2017). Activity and safety of crizotinib in patients with advanced clear-cell sarcoma with MET alterations: European Organization for Research and Treatment of Cancer phase II trial 90101 “CREATE.” Annals of Oncology, 28(12), 3000–3008. 10.1093/annonc/mdx527

Silk, A. D., Zasadil, L. M., Holland, A. J., Vitre, B., Cleveland, D. W., & Weaver, B. A. (2013). Chromosome missegregation rate predicts whether aneuploidy will promote or suppress tumors. Proceedings of the National Academy of Sciences of the United States of America, 110(44). 10.1073/pnas.1317042110

Simón-Carrasco, L., Pietrini, E., & López-Contreras, A. J. (2024). Integrated analysis of FHIT gene alterations in cancer. In Cell Cycle (Vol. 23, Number 1, pp. 92–113). Taylor and Francis Ltd. 10.1080/15384101.2024.2304509

Snyder, A., Makarov, V., Merghoub, T., Yuan, J., Zaretsky, J. M., Desrichard, A., Walsh, L. A., Postow, M. A., Wong, P., Ho, T. S., Hollmann, T. J., Bruggeman, C., Kannan, K., Li, Y., Elipenahli, C., Liu, C., Harbison, C. T., Wang, L., Ribas, A., … Chan, T. A. (2014). Genetic Basis for Clinical Response to CTLA-4 Blockade in Melanoma. New England Journal of Medicine, 371(23), 2189–2199. 10.1056/nejmoa1406498

Steele, C. D., Tarabichi, M., Oukrif, D., Webster, A. P., Ye, H., Fittall, M., Lombard, P., Martincorena, I., Tarpey, P. S., Collord, G., Haase, K., Strauss, S. J., Berisha, F., Vaikkinen, H., Dhami, P., Jansen, M., Behjati, S., Amary, M. F., Tirabosco, R., … Pillay, N. (2019). Undifferentiated Sarcomas Develop through Distinct Evolutionary Pathways. Cancer Cell, 35(3), 441–456.e8. 10.1016/j.ccell.2019.02.002

Su, X. A., Ma, D., Parsons, J. V., Replogle, J. M., Amatruda, J. F., Whittaker, C. A., Stegmaier, K., & Amon, A. (2021). RAD21 is a driver of chromosome 8 gain in Ewing sarcoma to mitigate replication stress. Genes and Development, 35(78), 556–572. 10.1101/gad.345454.120

Su, X. A., Stopsack, K. H., Schmidt, D. R., Ma, D., Li, Z., Scheet, P. A., Penney, K. L., Lotan, T. L., Abida, W., DeArment, E. G., Lu, K., Janas, T., Hu, S., Vander Heiden, M. G., Loda, M., Boselli, M., Amon, A., & Mucci, L. A. (2024). RAD21 promotes oncogenesis and lethal progression of prostate cancer. Proceedings of the National Academy of Sciences of the United States of America, 121(36). 10.1073/pnas.2405543121

Taylor, A. M., Shih, J., Ha, G., Gao, G. F., Zhang, X., Berger, A. C., Schumacher, S. E., Wang, C., Hu, H., Liu, J., Lazar, A. J., Caesar-Johnson, S. J., Demchok, J. A., Felau, I., Kasapi, M., Ferguson, M. L., Hutter, C. M., Sofia, H. J., Tarnuzzer, R., … Meyerson, M. (2018). Genomic and Functional Approaches to Understanding Cancer Aneuploidy. Cancer Cell, 33(4), 676–689.e3. 10.1016/j.ccell.2018.03.007

The National Genomic Research Library. (2024). Genomics England. 10.6084/m9.figshare.4530893

Uddin, I., Joshi, K., Oakes, T., Heather, J. M., Swanton, C., & Chain, B. (2019). An Economical, Quantitative, and Robust Protocol for High-Throughput T Cell Receptor Sequencing from Tumor or Blood (pp. 15–42). 10.1007/978-1-4939-8885-3_2

Umbreit, N. T., Zhang, C.-Z., Lynch, L. D., Blaine, L. J., Cheng, A. M., Tourdot, R., Sun, L., Almubarak, H. F., Judge, K., Mitchell, T. J., Spektor, A., & Pellman, D. (2020). MECHANISMS GENERATING CANCER GENOME COMPLEXITY FROM A SINGLE CELL DIVISION ERROR HHS Public Access. Science, 368(6488). 10.5061/dryad.rn8pk0p61

Van Loo, P., Nordgard, S. H., Lingjærde, O. C., Russnes, H. G., Rye, I. H., Sun, W., Weigman, V. J., Marynen, P., Zetterberg, A., Naume, B., Perou, C. M., Børresen-Dale, A. L., & Kristensen, V. N. (2010). Allele-specific copy number analysis of tumors. Proceedings of the National Academy of Sciences of the United States of America, 107(39), 16910–16915. 10.1073/pnas.1009843107

Wada, K., Sato, M., Araki, N., Kumeta, M., Hirai, Y., Takeyasu, K., Furukawa, K., & Horigome, T. (2014). Dynamics of WD-repeat containing proteins in SSU processome components. Biochemistry and Cell Biology, 92(3), 191–199. 10.1139/bcb-2014-0007

Wang, H. Y., Cogua, L. M., Tupper, C. J., & Silberstein, P. T. (2025). Clear Cell Sarcoma Incidence and Survival: A Surveillance, Epidemiology, and End Results (SEER) Database Analysis. Cureus. 10.7759/cureus.85606

Wang, J., & Laframboise, T. (2019). CytoConverter: A web-based tool to convert karyotypes to genomic coordinates. BMC Bioinformatics, 20(1). 10.1186/s12859-019-3062-4

Waters, C. E., Saldivar, J. C., Hosseini, S. A., & Huebner, K. (2014). The FHIT gene product: Tumor suppressor and genome “caretaker.” In Cellular and Molecular Life Sciences (Vol. 71, Number 23, pp. 4577–4587). Birkhauser Verlag AG. 10.1007/s00018-014-1722-0

WHO Classification of Tumours Editorial Board. (2026). Soft tissue and bone tumours [Internet; beta version ahead of print] (6th ed., Vol. 3). International Agency for Research on Cancer. https://tumourclassification.iarc.who.int/chapters/87

Woodfield, S. E., Shi, Y., Patel, R. H., Chen, Z., Shah, A. P., Whitlock, R. S., Ibarra, A. M., Larson, S. R., Sarabia, S. F., Badachhape, A., Starosolski, Z., Ghaghada, K. B., Sumazin, P., Annis, D. A., López-Terrada, D., & Vasudevan, S. A. (2021). MDM4 inhibition: a novel therapeutic strategy to reactivate p53 in hepatoblastoma. Scientific Reports, 11(1). 10.1038/s41598-021-82542-4

Yoshihara, K., Shahmoradgoli, M., Martínez, E., Vegesna, R., Kim, H., Torres-Garcia, W., Treviño, V., Shen, H., Laird, P. W., Levine, D. A., Carter, S. L., Getz, G., Stemke-Hale, K., Mills, G. B., & Verhaak, R. G. W. (2013). Inferring tumour purity and stromal and immune cell admixture from expression data. Nature Communications, 4. 10.1038/ncomms3612

Yu, G., Wang, L. G., Han, Y., & He, Q. Y. (2012). ClusterProfiler: An R package for comparing biological themes among gene clusters. OMICS A Journal of Integrative Biology, 16(5), 284–287. 10.1089/omi.2011.0118

Zhang, C., Yin, C., Wang, L., Zhang, S., Qian, Y., Ma, J., Zhang, Z., Xu, Y., & Liu, S. (2014). HSPC111 governs breast cancer growth by regulating ribosomal biogenesis. Molecular Cancer Research, 12(4), 583–594. 10.1158/1541-7786.MCR-13-0168

Zhang, Y., Chen, Y., Yang, C., Seger, N., Hesla, A. C., Tsagkozis, P., Larsson, O., Lin, Y., & Haglund, F. (2021). TERT promoter mutation is an objective clinical marker for disease progression in chondrosarcoma. Modern Pathology, 34(11), 2020–2027. 10.1038/s41379-021-00848-0

Zheng, S. C., Webster, A. P., Dong, D., Feber, A., Graham, D. G., Sullivan, R., Jevons, S., Lovat, L. B., Beck, S., Widschwendter, M., & Teschendorff, A. E. (2018). A novel cell-type deconvolution algorithm reveals substantial contamination by immune cells in saliva, buccal and cervix. Epigenomics, 10(7), 925–940. 10.2217/epi-2018-0037

